# Head-to-Head Comparison of Flortaucipir and MK6240 tau-PET to Detect Subtle Objective Cognitive Impairment in Clinically Unimpaired Individuals

**DOI:** 10.64898/2026.11.01.26367030

**Authors:** Arthur C. Macedo, Étienne Aumont, Lydia Trudel, Seyyed Ali Hosseini, Joseph Therriault, Gleb Bezgin, Aurélie Lebrun, Delphine Oliva-Lopez, Firoza Z Lussier, Tevy Chan, Cécile Tissot, Nesrine Rahmouni, Brandon Hall, Isabella Bizzi, Cecilia Boccalini, Juliana Beker Godoy, Guilherme Povala, Pamela C. L. Ferreira, Bruna Bellaver, Jaime Fernandez-Arias, Livia Amaral, Emma Ruppert, Marina Scop Medeiros, Rayan Mroué, Andréia Rocha, Maxime Montembeault, Belen Pascual, Joseph Masdeu, Brian A Gordon, Val J. Lowe, Hwamee Oh, David N. Soleimani Meigooni, Juan Fortea, Suzanne Baker, Tharick A Pascoal, Pedro Rosa-Neto, the HEAD Study

## Abstract

**Background and Objectives:** Tau-PET is associated with subtle objective cognitive impairment (SOCI) in clinically unimpaired (CU) individuals, but whether the spatial patterns and strengths of these associations differ across tracers remains unclear. We performed a head-to-head comparison of [¹□F]MK6240 and [¹□F]flortaucipir to investigate their associations with SOCI in CU individuals.

**Methods:** We cross-sectionally assessed CU participants from the HEAD study who underwent both tau-PET scans. Linear regressions examined associations between domain-specific cognition and tau burden using standardized uptake value ratios (SUVR) in regional and voxelwise analyses, alongside regional spatial extent of tauopathy (SEOT) measures. Tau burden was compared across groups representing different levels of SOCI. Models adjusted for age, sex, and education, with additional adjustment for amyloid-β burden. Centiloid-stratified analyses examined associations across amyloid-β burden levels.

**Results:** Among 297 participants (mean [SD] age, 67.0 [8.4] years; 179 [60.3%] female), higher [¹□F]MK6240 SUVR was associated with poorer memory performance. Greater SEOT with either tracer was associated with poorer memory and showed nominal associations with executive dysfunction. Regional association strengths did not differ significantly between tracers, while SEOT yielded comparable or stronger associations than SUVR. Centiloid-stratified analyses revealed tau– cognition associations primarily at intermediate and high amyloid-β burden. [¹□F]MK6240 showed more spatially extensive voxelwise associations with memory and across SOCI group comparisons. Regional analyses also yielded more statistically significant differences across SOCI groups with [¹□F]MK6240 than with flortaucipir. These differences were predominantly driven by participants meeting actuarial mild cognitive impairment criteria, who exhibited greater tau burden than groups with less pronounced psychometric impairment. Across tracers, SEOT yielded more consistently significant differences among SOCI groups than SUVR.

**Discussion:** Both tracers showed associations with SOCI in CU individuals, with broadly comparable regional association strengths despite differences in the spatial extent and statistical significance of their findings. SEOT complemented SUVR by identifying additional significant associations and group differences. These findings suggest that tracer choice and quantification approach may influence the spatial distribution and statistical significance of observed tau–cognition relationships without necessarily altering their overall strength.

## Introduction

In the revised Alzheimer’s Association research and diagnostic framework, “clinical stage 2 - transitional decline” is recognized as a phase of Alzheimer’s disease (AD), encompassing subtle cognitive or behavioral changes with minimal impact on daily functioning.^1^ Among these manifestations are objectively detectable cognitive changes that do not meet criteria for mild cognitive impairment (MCI) or dementia, a phenotype broadly referred to in the literature as subtle objective cognitive impairment (SOCI).^2^ SOCI is identified through standardized neuropsychological assessment and may occur with or without subjective cognitive complaints.^2^ However, the specific criteria used to operationalize SOCI vary widely and include performance on individual tests or composite scores, longitudinal intra-individual decline, clinical ratings, and data-driven approaches.^2^ Although these definitions converge on the detection of subtle cognitive changes before conventional MCI criteria are met, associations between SOCI and AD biomarkers have varied.^3–6^ This variability may reflect not only differences in SOCI definitions but also differences in the sensitivity of biomarkers and measurement approaches.

Among currently available biomarkers of core AD pathologies, tau positron emission tomography (tau-PET) is most closely associated with cognitive symptoms.^7^ In clinically unimpaired (CU) individuals, greater tau-PET uptake has been associated with poorer performance across several cognitive domains.^8–11^ Understanding these early associations has become increasingly relevant as tau-PET is incorporated as an endpoint in preventive trials.^12^ In this context, tracer differences in sensitivity could influence the correspondence between tau-PET and clinical outcomes, affecting the interpretation and comparability of biomarker findings across trials. However, it remains unclear whether newer, more sensitive second-generation tau-PET tracers,^13^ and spatial extent metrics that may better reflect the clinical expression of AD,^14,15^ provide stronger associations with SOCI.

Therefore, we compared the FDA-approved [¹□F]Flortaucipir and [¹□F]MK6240 head-to-head regarding their associations with subtle cognitive differences in preclinical AD. Specifically, we aimed to clarify how tau-PET tracer choice influences the characterization of the earliest measurable cognitive manifestations of AD. Secondarily, we investigated whether tau spatial extent provides information beyond conventional SUVR measures in capturing the tau-PET correlates of SOCI.

## Methods

### Participants

In this cross-sectional study, we assessed individuals diagnosed as CU from the HEAD study (https://head-study.info/), recruited across eight sites in the United States and Canada between November 2022 and March 2025. CU status was defined as not meeting standard diagnostic criteria for MCI or dementia due to AD or another cause, and having a global Clinical Dementia Rating (CDR) score of 0.^13,16,17^ Diagnostic ascertainment was performed by multidisciplinary teams of specialists in dementia, following comprehensive clinical examinations.

Inclusion criteria were age ≥50 years, an available study partner to assess functioning, ability, and willingness to undergo MRI and PET, and fluency in a coordinating-centre-approved language.^13^ Exclusions were inability to consent, pregnancy/breastfeeding, and conditions or medications compromising procedural safety or tolerability.^13^ No *a priori* sample-size calculation was performed because all participants meeting the inclusion criteria with complete, quality-controlled data for all study variables were included.

### Imaging acquisition and processing

Participants underwent structural MRI, tau-PET imaging with both [¹□F]Flortaucipir and [¹□F]MK6240, and amyloid-β (Aβ) PET imaging (either [¹¹C]PiB, [¹□F]AZD4694, or [¹□F]Florbetaben, depending on site preference).

HEAD imaging protocols have been described previously.^13^ Briefly, MRI was acquired on 3T scanners using ADNI-4–aligned protocols,^18^ including high-resolution sagittal T1-weighted images obtained with three-dimensional accelerated MPRAGE or IRSPGR sequences. PET acquisition windows after intravenous bolus injection were 40–60 or 50–70 min for [¹¹C]PiB, 90–110 min for [¹□F]florbetaben, 50–70 min for [¹□F]AZD4694, 90–110 min for [¹□F]MK6240, and 80–100 min for [¹□F]flortaucipir. PET images were reconstructed into four 5-min frames using scanner-specific ordered-subset expectation maximization–based algorithms with standardized parameters and no smoothing, with corrections for attenuation, motion, dead time, radioactive decay, scatter, and random events.

Images were processed using a validated PET–MRI pipeline.^13^ T1-weighted MRI was nonlinearly registered to the ICBM 2020 Nonlinear Symmetric template using Advanced Normalization Tools (ANTs).^19^ PET images were linearly co-registered to native-space T1-weighted MRI, then warped into ICBM 2020 stereotaxic space using the concatenated PET-to-T1 and T1-to-template transformations. T1-derived whole-brain masks excluded extracerebral voxels, and scanner-specific Gaussian smoothing yielded a uniform 8-mm full width at half maximum.

### PET Image Quantification and Analysis

Tau-PET standardized uptake value ratio (SUVR) images were derived using inferior cerebellar gray matter as the reference region.^13^ Regional mean SUVR values were computed to quantify tau load. Using the same regions-of-interest (ROIs), we also derived the spatial extent of tauopathy (SEOT), defined as the proportion of voxels exceeding 2.5 standard deviations above the mean of a reference group of 27 CU young individuals (mean [SD] age 22.7 [1.8] years; 59% females) scanned head-to-head with both tau-PET tracers.^20^

Tau-PET metrics were extracted from previously described medial temporal lobe (MTL) and neocortical ROIs representing successive phases of AD-related tau propagation (**Supplementary Figures 1A-B**).^14^ The MTL mask included the transentorhinal, entorhinal, amygdala, parahippocampal gyrus, perirhinal, and hippocampus CA1.^14,21^ The neocortical mask consisted of the inferior, lateral, and superior temporal, inferior and superior parietal, supramarginal, insula, anterior and posterior cingulate, precuneus, cuneus, lateral occipital, pericalcarine, inferior and superior frontal, orbitofrontal, rostromedial frontal, paracentral, postcentral, and precentral regions.^14^ We additionally examined the entorhinal cortex as an individual ROI because it is commonly considered among the earliest regions affected in tau-PET staging frameworks.^22^ To mitigate off-target binding,^23,24^ cortical ROIs were eroded to reduce [¹□F]MK6240 meningeal spill-in,^24^ and hippocampal analyses were restricted to CA1 to reduce [¹□F]flortaucipir choroid plexus contamination.^21,23^

Global Aβ SUVR was calculated in a neocortical composite using whole cerebellum as reference and converted to Centiloids (CL).^25^ Aβ burden was categorized according to Amyloid Imaging to Prevent Alzheimer’s Disease (AMYPAD)-recommended CL ranges as low (<10 CL; CL_LOW_), intermediate (10–30 CL; CL_INT_), or high (>30 CL; CL_HIGH_).^26^

### Cognitive assessment

Neuropsychological assessment followed the National Alzheimer’s Coordinating Center Uniform Data Set version 3.0 (UDSv3),^27^ from which four validated domain-specific composites were calculated.^28^ The memory composite consisted of the Craft Story Immediate and Delayed Recall scores (paraphrase scoring) and the Benson Complex Figure delayed-recall score. Language comprised category fluency for animals and vegetables, the Multilingual Naming Test (MINT) total score, and phonemic fluency for F- and L-words. Attention comprised the Trail Making Test (TMT) Part A completion time (reverse-scored) and Forward Number Span correct trials. Executive function comprised the TMT Part B completion time (reverse-scored) and Backward Number Span correct trials.

### SOCI psychometric staging

We classified participants into SOCI psychometric stages by adapting actuarial criteria for objectively defined subtle cognitive decline (obj-SCD)^29,30^ and MCI^31^ to the UDSv3 battery. These criteria identify abnormal performance across memory, language, and attention/executive function. The original criteria were adapted using corresponding UDSv3 measures for each domain (**Figure 1A**): Craft Story delayed recall (paraphrase scoring) and Benson Figure delayed recall for memory; Animal Fluency and MINT for language; and TMT A and B for attention/executive function. Individual test performance was considered abnormal at ≥1 SD below demographically expected values using published UDSv3 norms accounting for age, sex, and education.^27,29–31^

**Figure 1.**
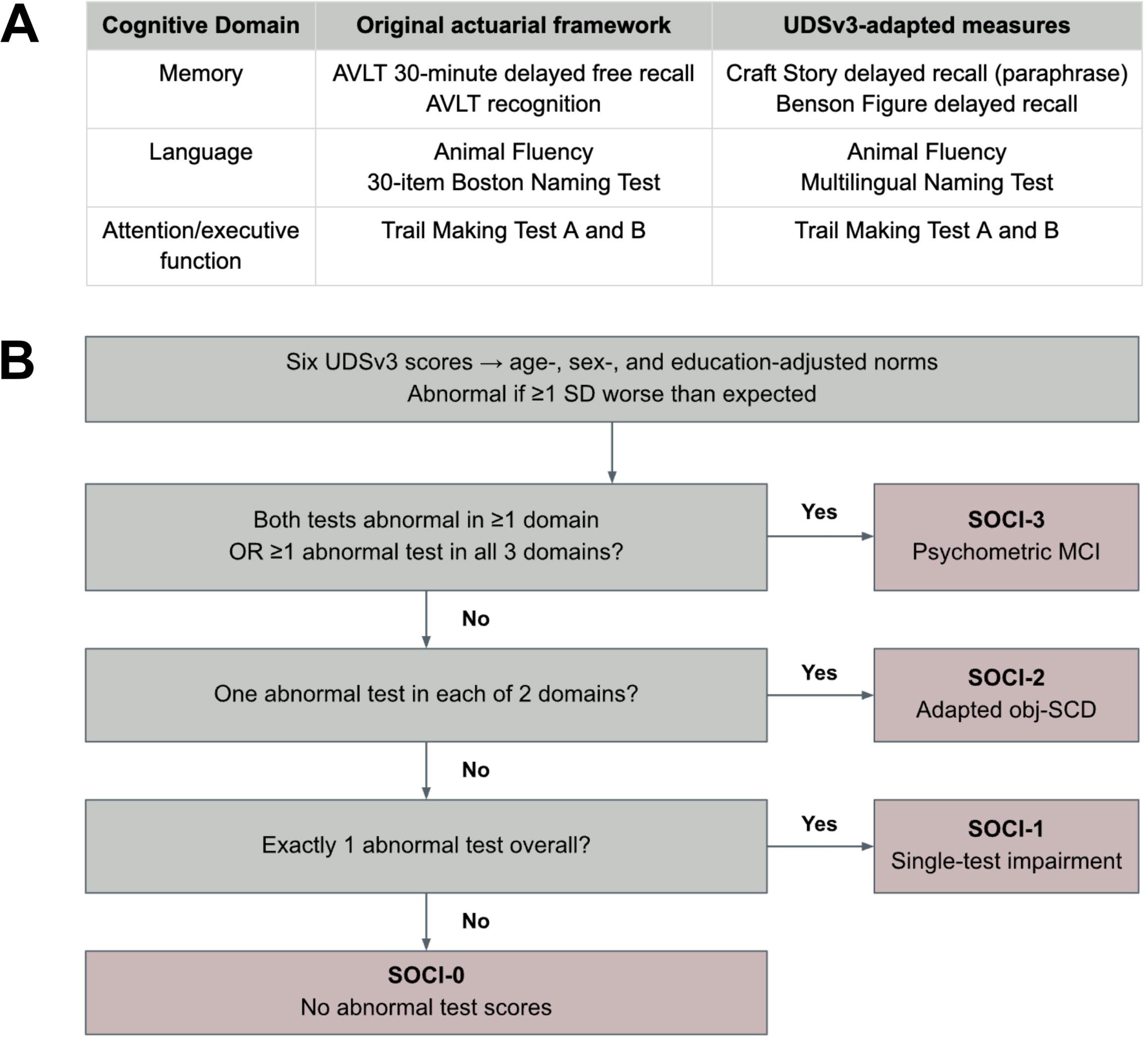
Neuropsychological measures and hierarchical algorithm for SOCI staging. **(A)** Original actuarial measures and their UDSv3 adaptations. AVLT process scores and the FAQ were excluded from the adapted criteria. **(B)** Hierarchical assignment to four mutually exclusive SOCI stages. Abnormal performance was defined as ≥1 SD worse than expected using UDSv3 norms adjusted for age, sex, and education. The algorithm was applied to clinically unimpaired participants with a CDR = 0. AVLT, Rey Auditory Verbal Learning Test; CDR, clinical dementia rating; FAQ, Functional Activities Questionnaire; MCI, mild cognitive impairment; obj-SCD, objectively defined subtle cognitive decline; SD, standard deviation; UDSv3, Uniform Data Set version 3.

Participants were assigned hierarchically to four mutually exclusive SOCI stages based on abnormal scores within and across domains (**Figure 1B**). Those with two abnormal tests within one domain or one abnormal test in each of three domains met actuarial MCI criteria³¹ and were classified as SOCI-3. They were retained within the SOCI framework because they lacked a clinical MCI diagnosis based on medical assessment and CDR scores, consistent with previous pre-MCI neuropsychological definitions of SOCI.^2^ Among remaining participants, those with one abnormal test in each of two domains met adapted obj-SCD criteria and were classified as SOCI-2.^29,30^ One abnormal test defined SOCI-1 (single-test impairment), and no abnormal tests defined SOCI-0 (no impairment).

Thomas et al. extended Edmonds’ criteria to include word-list process scores, including intrusion errors and learning slope;^29,30^ these were unavailable in UDSv3 and omitted from SOCI-2 definition. The Functional Activities Questionnaire was excluded from SOCI staging, consistent with more recent operationalizations of actuarial criteria.^2^

We also explored a simplified SOCI definition using the Montreal Cognitive Assessment (MoCA).^32^ Consistent with previous research, performance >1 z score below age-, sex-, and education-adjusted expectations from UDSv3 norms was considered subtly abnormal.^33^

### Statistical analyses

Statistical analyses were conducted in R version 4.4.1. ROI-based associations between tau-PET SUVR and cognitive performance were evaluated using separate linear regression models for each cognitive domain, tracer, and ROI (MTL and neocortex). Cognitive scores and tau-PET measures were z-standardized using the full study sample. Models were adjusted for age, sex, and education, with additional models further adjusted for CL. In secondary analyses, those models were repeated using tau-PET SEOT in place of SUVR as the main predictor. False discovery rate (FDR) correction was performed using the Benjamini–Hochberg procedure separately for each tau-PET metric-cognitive outcome pair, with each family comprising four tests (2 tau-PET tracers × 2 ROIs). Sensitivity analyses were conducted using the entorhinal as the ROI.

Effect-size differences were evaluated using 5,000 paired participant-level bootstrap resamples of 297 participants sampled with replacement. Models used identical resampled participants, outcomes, and covariates; standardized coefficients were recalculated to obtain paired tracer ([¹□F]MK6240 minus [¹□F]flortaucipir) and metric (SEOT minus SUVR) differences.

We further explored SUVR-cognition associations within CL-defined subgroups using linear regressions adjusting only for demographic covariates. FDR correction was applied separately for each tau-PET ROI-cognitive domain pair across the three CL groups and two tau-PET tracers, yielding six tests per correction family.

Voxel-based linear regression analyses were performed in R (*RMINC* package), with tau-PET as the dependent variable and cognitive measures as the independent variables, adjusting for age, sex, and education. Secondary models further adjusted for CL. Statistical significance was determined using an FDR p<0.05. Analyses were restricted to a mask encompassing all Braak regions defined in the CerebrA atlas (**Supplementary Figure 1C**).^34^

Differences in regional tau-PET measures across SOCI stages were evaluated using analysis of covariance (ANCOVA) adjusted for age, sex, and education. Secondary models were additionally adjusted for CL. Estimated marginal means (EMM) were compared using pairwise t-test with Tukey adjustment. Voxelwise differences in tau-PET across SOCI stages were assessed using linear regression in *RMINC*, adjusting for the same covariates. SOCI stages were modelled as a four-level categorical variable, with SOCI-0 as the reference and statistical significance defined at FDR p<0.05.

### Standard Protocol Approvals, Registrations, and Patient Consents

The study was approved by the central institutional review board, Advarra (United States), and the Douglas Research Institute Research Ethics Board (Canada). Written informed consent was obtained from all participants.

### Data Availability

Requests for raw or analysed data and materials should be directed to the corresponding author and are subject to investigator and institutional review for intellectual property and confidentiality restrictions. Anonymised data and materials will be shared with qualified academic investigators to replicate this Article’s procedures and results under a material transfer agreement.

### AI Use

The authors used ChatGPT-6.1 to improve clarity and readability, reviewed and approved all suggested revisions, and retain full responsibility for the manuscript’s content.

## Results

We studied 297 participants with a mean (SD) age of 67.0 (8.4) years, of whom 179 (60.3%) were female (**Table 1**). Overall, 146 (49.2%) participants were classified as SOCI-0, 94 (31.6%) as SOCI-1, 37 (12.5%) as SOCI-2, and 20 (6.7%) as SOCI-3. The groups did not differ in age, sex, education, or CL values (all p>0.05). MoCA scores differed across groups (p=0.0002); compared with SOCI-0 participants, scores were lower in the SOCI-2 and SOCI-3 groups, and the SOCI-3 group had lower scores than the SOCI-1 group (Holm-adjusted p<0.05).

**Table 1.**
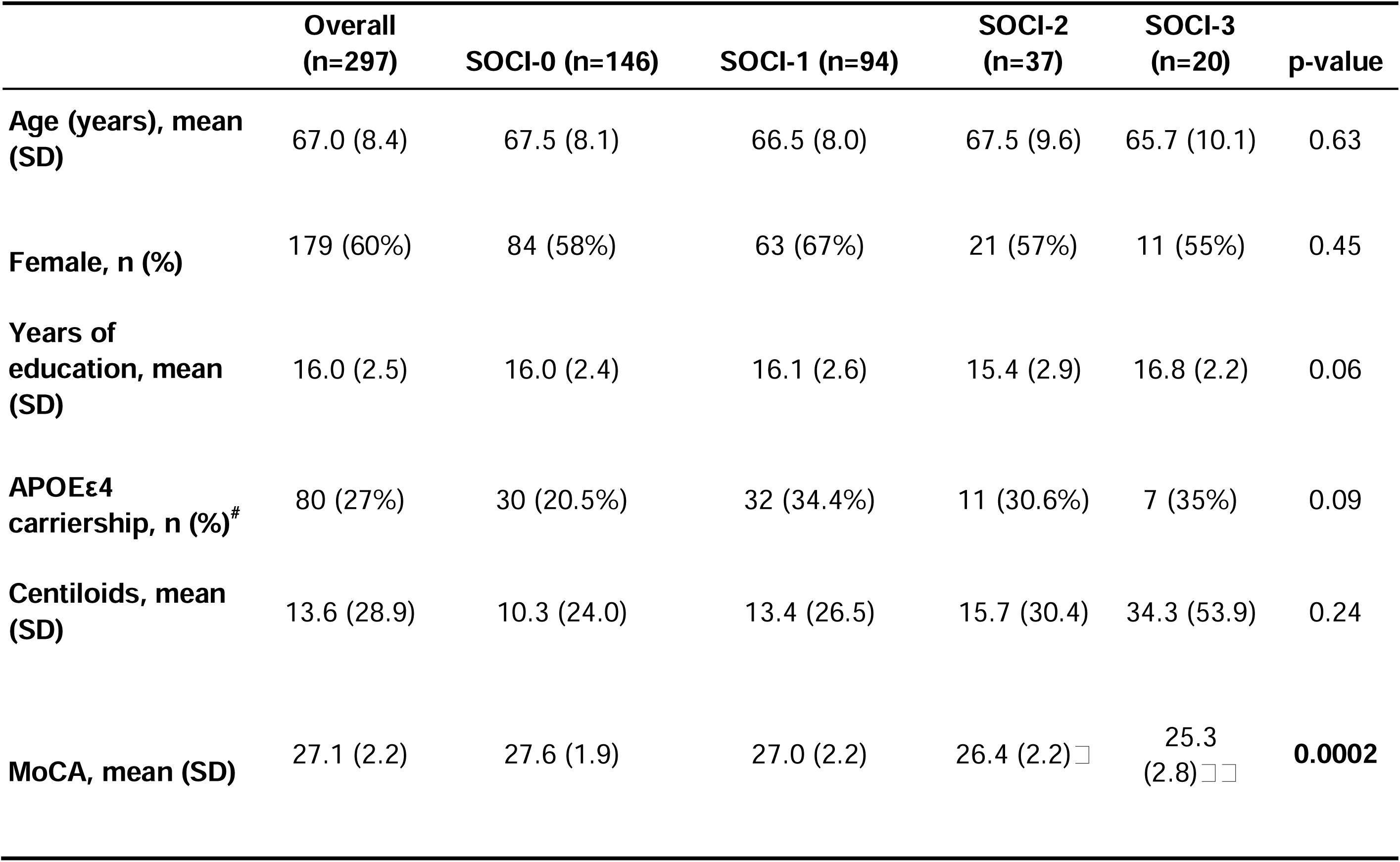
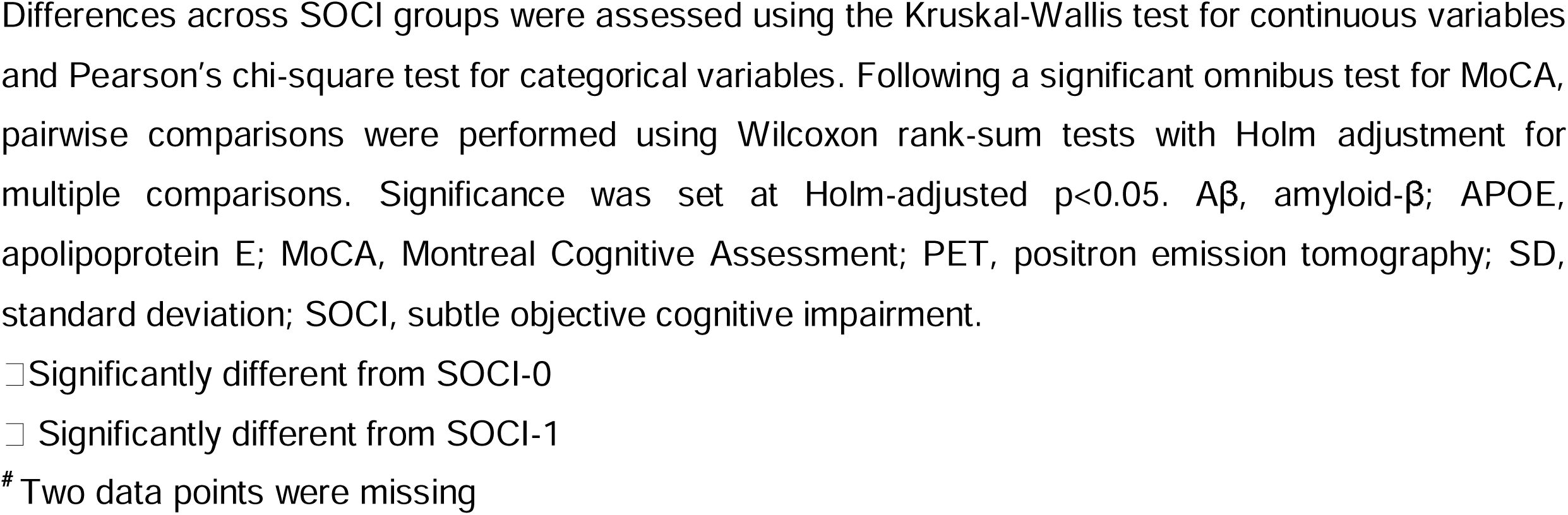
Demographic, clinical, and biomarker characteristics of the participants.

|  | **Overall (n=297)** | **SOCI-0 (n=146)** | **SOCI-1 (n=94)** | **SOCI-2 (n=37)** | **SOCI-3 (n=20)** | **p-value** |
|---|---|---|---|---|---|---|
| **Age (years), mean (SD)** | 67.0 (8.4) | 67.5 (8.1) | 66.5 (8.0) | 67.5 (9.6) | 65.7 (10.1) | 0.63 |
| **Female, n (%)** | 179 (60%) | 84 (58%) | 63 (67%) | 21 (57%) | 11 (55%) | 0.45 |
| **Years of education, mean (SD)** | 16.0 (2.5) | 16.0 (2.4) | 16.1 (2.6) | 15.4 (2.9) | 16.8 (2.2) | 0.06 |
| **APOEε4 carriership, n (%)**<sup>#</sup> | 80 (27%) | 30 (20.5%) | 32 (34.4%) | 11 (30.6%) | 7 (35%) | 0.09 |
| **Centiloids, mean (SD)** | 13.6 (28.9) | 10.3 (24.0) | 13.4 (26.5) | 15.7 (30.4) | 34.3 (53.9) | 0.24 |
| **MoCA, mean (SD)** | 27.1 (2.2) | 27.6 (1.9) | 27.0 (2.2) | 26.4 (2.2)□ | 25.3 (2.8)□□ | **0.0002** |

Differences across SOCI groups were assessed using the Kruskal-Wallis test for continuous variables and Pearson’s chi-square test for categorical variables. Following a significant omnibus test for MoCA, pairwise comparisons were performed using Wilcoxon rank-sum tests with Holm adjustment for multiple comparisons. Significance was set at Holm-adjusted p<0.05. Aβ, amyloid-β; APOE, apolipoprotein E; MoCA, Montreal Cognitive Assessment; PET, positron emission tomography; SD, standard deviation; SOCI, subtle objective cognitive impairment.
□Significantly different from SOCI-0
□ Significantly different from SOCI-1
<sup>#</sup> Two data points were missing

### ROI-based associations between tau-PET SUVR and cognitive composites

After adjustment for age, sex, and education, greater [¹⁸F]MK6240 SUVR was associated with poorer memory in the MTL (β=−0.15, 95% CI −0.27 to −0.03, p_FDR_=0.04) and neocortex (β=−0.13, 95% CI −0.25 to −0.02, p_FDR_=0.04), whereas [¹□F]flortaucipir SUVR was not (**Figure 2A-B; Supplementary Table 1**). Neither tracer showed significant associations with language, attention, or executive function. Paired bootstrap comparisons showed nominally more negative standardized MTL SUVR coefficients for [¹□F]MK6240 than [¹□F]flortaucipir for memory (p=0.046) and language (p=0.03), with no other nominal differences; none survived FDR correction (**Figure 2C**). After CL adjustment, [¹□F]MK6240–memory associations attenuated, and no SUVR-cognition associations were significant (**Supplementary Table 2**).

**Figure 2.**
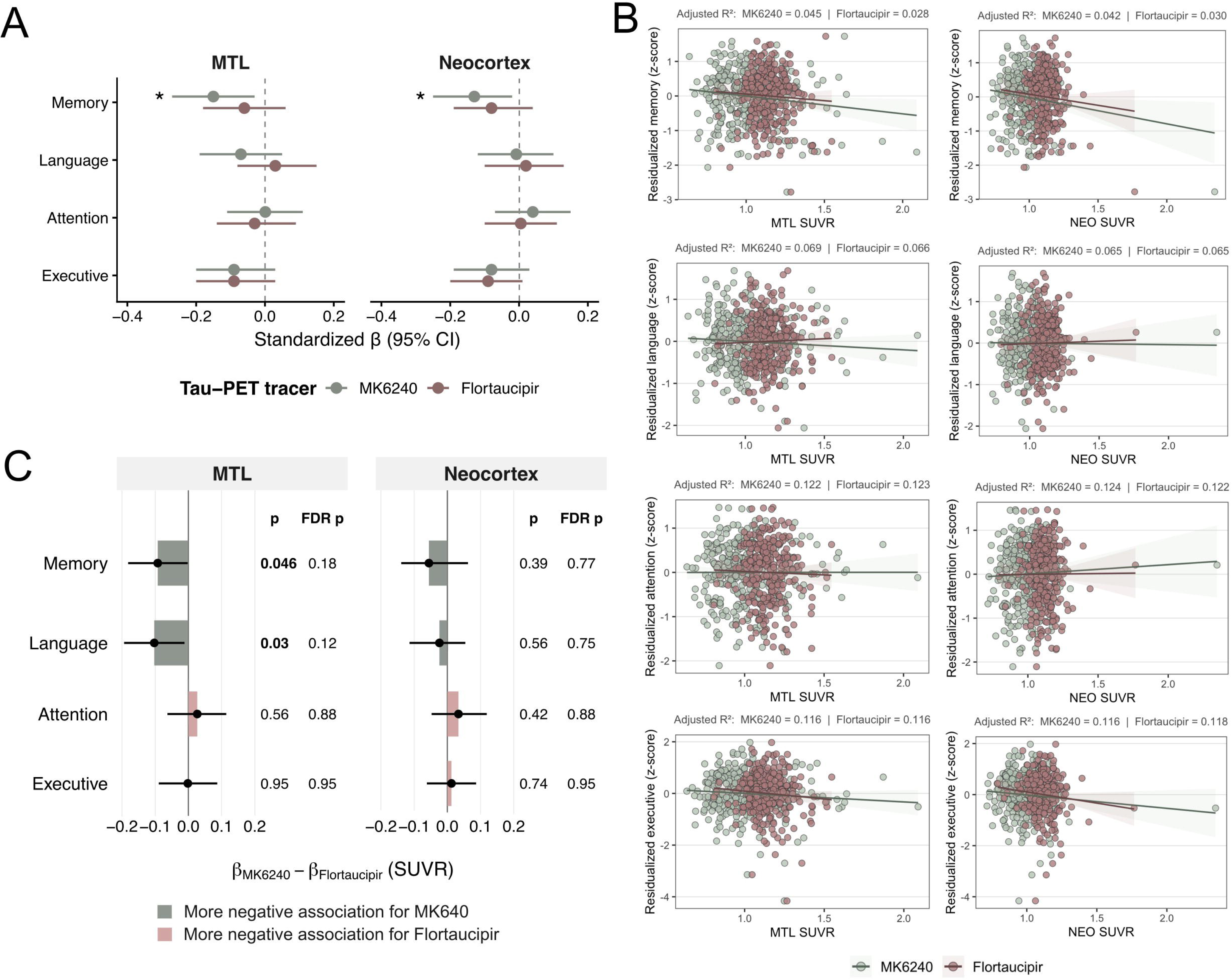
Associations between regional tau-PET load and cognitive performance. **(A)** Standardised regression coefficients (β) and 95% confidence intervals for associations between MTL or neocortical tau-PET SUVR and cognitive performance. Linear models were adjusted for age, sex, and years of education. * indicates Benjamini-Hochberg-adjusted p_FDR_ < 0.05. FDR correction was applied across the four tracer-by-region associations within each cognitive domain. **(B)** Scatter plots of the associations presented in **A**. **(C)** Paired differences in standardised coefficients between tracers ([¹□F]MK6240 minus [¹□F]flortaucipir), estimated using 5,000 paired bootstrap resamples. Bolded values indicate statistically significant comparisons (p<0.05). MTL, medial temporal lobe; NEO, neocortex; SUVR, standardised uptake value ratio.

In sensitivity analyses using the entorhinal as the ROI, greater [¹⁸F]MK6240 SUVR was associated with poorer memory (β=−0.11, 95% CI −0.20 to −0.02, pFDR=0.03), whereas [¹⁸F]flortaucipir SUVR was not; neither tracer showed significant associations with other domains (**Supplementary Table 3**). CL adjustment attenuated the entorhinal [¹⁸F]MK6240-memory association (β=−0.06, 95% CI −0.16 to 0.04, p=0.22, p_FDR_=0.45) (**Supplementary Table 4**).

### ROI-based associations between tau-PET SEOT and cognitive composites

After adjustment for age, sex, and education, greater SEOT was associated with poorer memory for MTL [¹⁸F]MK6240 (β=−0.16, 95% CI −0.28 to −0.04, p_FDR_=0.01), neocortical [¹⁸F]MK6240 (β=−0.18, 95% CI −0.29 to −0.07, p_FDR_=0.003), MTL [¹⁸F]flortaucipir (β=−0.16, 95% CI −0.28 to −0.04, p_FDR_=0.01), and neocortical [¹⁸F]flortaucipir (β=−0.19, 95% CI −0.31 to −0.08, p_FDR_=0.003; **Figure 3A-B; Supplementary Table 5**).

**Figure 3.**
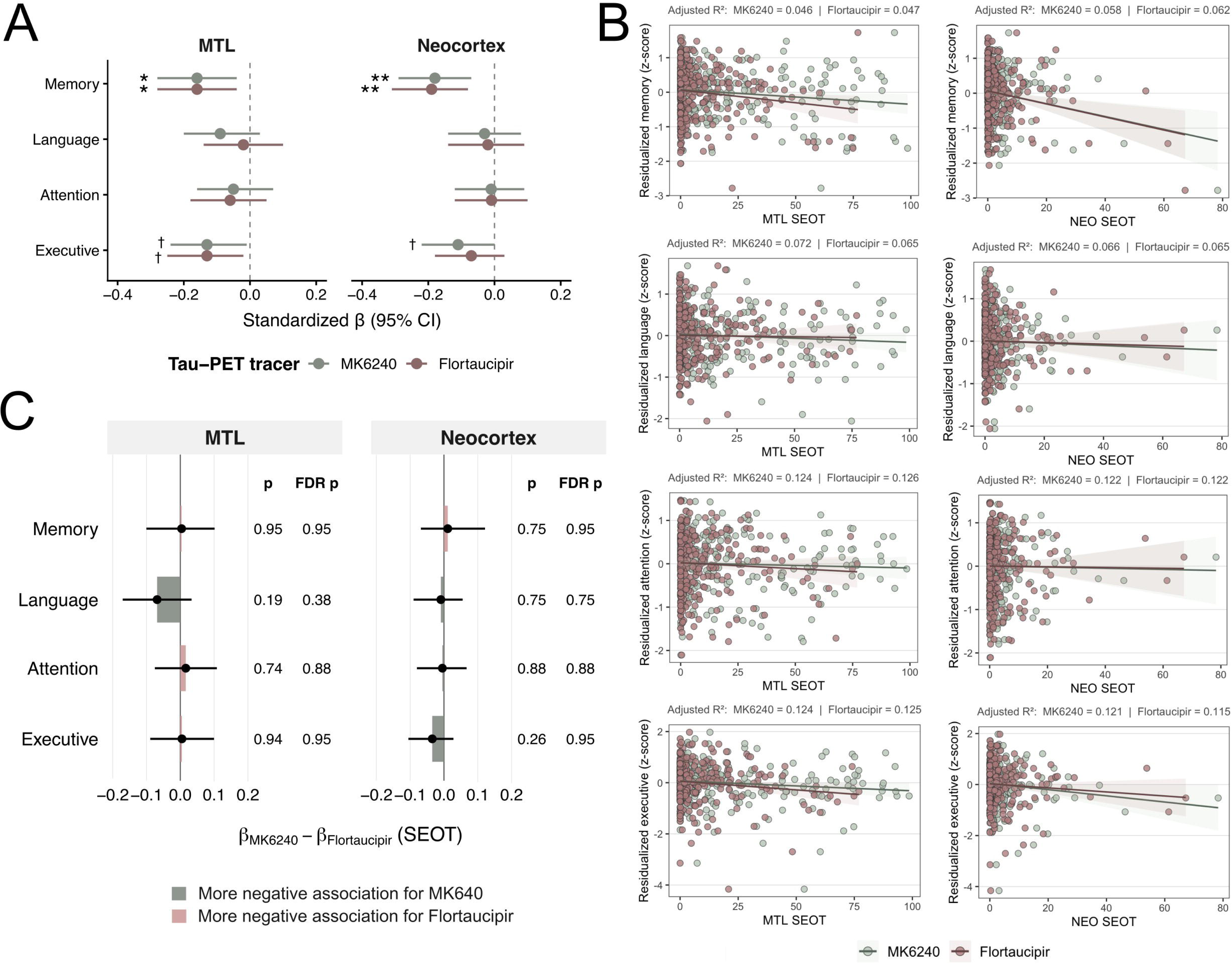
Associations between regional tau-PET spatial extent and cognitive performance. **(A)** Standardised regression coefficients (β) and 95% confidence intervals for associations between MTL or neocortical tau-PET SEOT and cognitive performance. Linear models were adjusted for age, sex, and years of education. † indicates nominal significance (p<0.05) that did not survive FDR correction; * and ** indicate Benjamini-Hochberg-adjusted p_FDR_<0.05 and p_FDR_<0.01, respectively. FDR correction was applied across the four tracer-by-region associations within each cognitive domain. **(B)** Scatter plots of the associations presented in **A**. **(C)** Paired differences in standardised coefficients between tracers ([¹□F]MK6240 minus [¹□F]flortaucipir), estimated using 5,000 paired bootstrap resamples. MTL, medial temporal lobe; NEO, neocortex; SEOT, spatial extent of tauopathy.

Poorer executive performance was nominally associated with MTL SEOT for [¹⁸F]MK6240 (β=−0.13, 95% CI −0.24 to −0.01, p=0.03) and [¹⁸F]flortaucipir (β=−0.13, 95% CI −0.25 to −0.02, p=0.02), and neocortical [¹⁸F]MK6240 SEOT (β=−0.11, 95% CI −0.22 to 0.00, p=0.048); none survived FDR correction. Neither tracer showed significant language or attention associations.

Paired bootstrap comparisons showed no between-tracer differences in standardized SEOT coefficients across domains or ROIs (**Figure 3C**). Within tracers, SEOT coefficients were more negative than SUVR coefficients for [¹□F]MK6240 MTL–attention (p_FDR_=0.03) and [¹□F]flortaucipir–memory associations in the MTL and neocortex (both p_FDR_=0.02; **Supplementary Figure 2**). Differences nominally favoured SEOT for [¹□F]MK6240 MTL–executive function (p=0.049, p_FDR_=0.20) and neocortex–attention associations (p=0.04, p_FDR_=0.08); no other differences were significant.

After CL adjustment, the neocortical SEOT–memory association remained significant for [¹⁸F]flortaucipir (β=−0.15, 95% CI −0.27 to −0.04, p_FDR_=0.03) but was only nominally significant for [¹⁸F]MK6240 (β=−0.13, 95% CI −0.25 to −0.01, p=0.03, p_FDR_=0.07; **Supplementary Table 6**). MTL SEOT-executive associations remained nominally significant for both tracers (both p_FDR_=0.077). No other associations were significant.

### Tau-PET Associations With Cognition Emerge at Intermediate and High Amyloid-β Burden

In exploratory CL-stratified analyses, higher MTL SUVR was nominally associated with poorer memory in CL_INT_ for [¹⁸F]flortaucipir (β=−0.38, 95% CI −0.67 to −0.08, p=0.01) and [¹⁸F]MK6240 (β=−0.34, 95% CI −0.64 to −0.04, p=0.03; **Figure 3**). Neocortical SUVR was nominally associated with poorer executive performance for both tracers in CL_INT_ and poorer memory for [¹□F]MK6240 in CL_HIGH_. No associations survived FDR correction or were nominally significant in CL_LOW_.

### [¹□F]MK6240 and [¹□F]Flortaucipir show differential topographic voxelwise associations with cognitive performance

After adjustment for age, sex, and education, voxelwise memory associations were more extensive for [¹□F]MK6240 than [¹□F]flortaucipir, encompassing 17.1% versus 0.7% of Braak-region voxels (**Figure 4A-B**). [¹□F]MK6240 showed broader neocortical clusters and additional MTL clusters, whereas no MTL clusters were observed for [¹□F]flortaucipir, consistent with the ROI-based findings. Only [¹□F]flortaucipir showed significant attention associations, mainly in a small bilateral occipital cluster (0.3% of voxels). Neither tracer showed significant language or executive associations, consistent with ROI-based SUVR findings.

**Figure 4.**
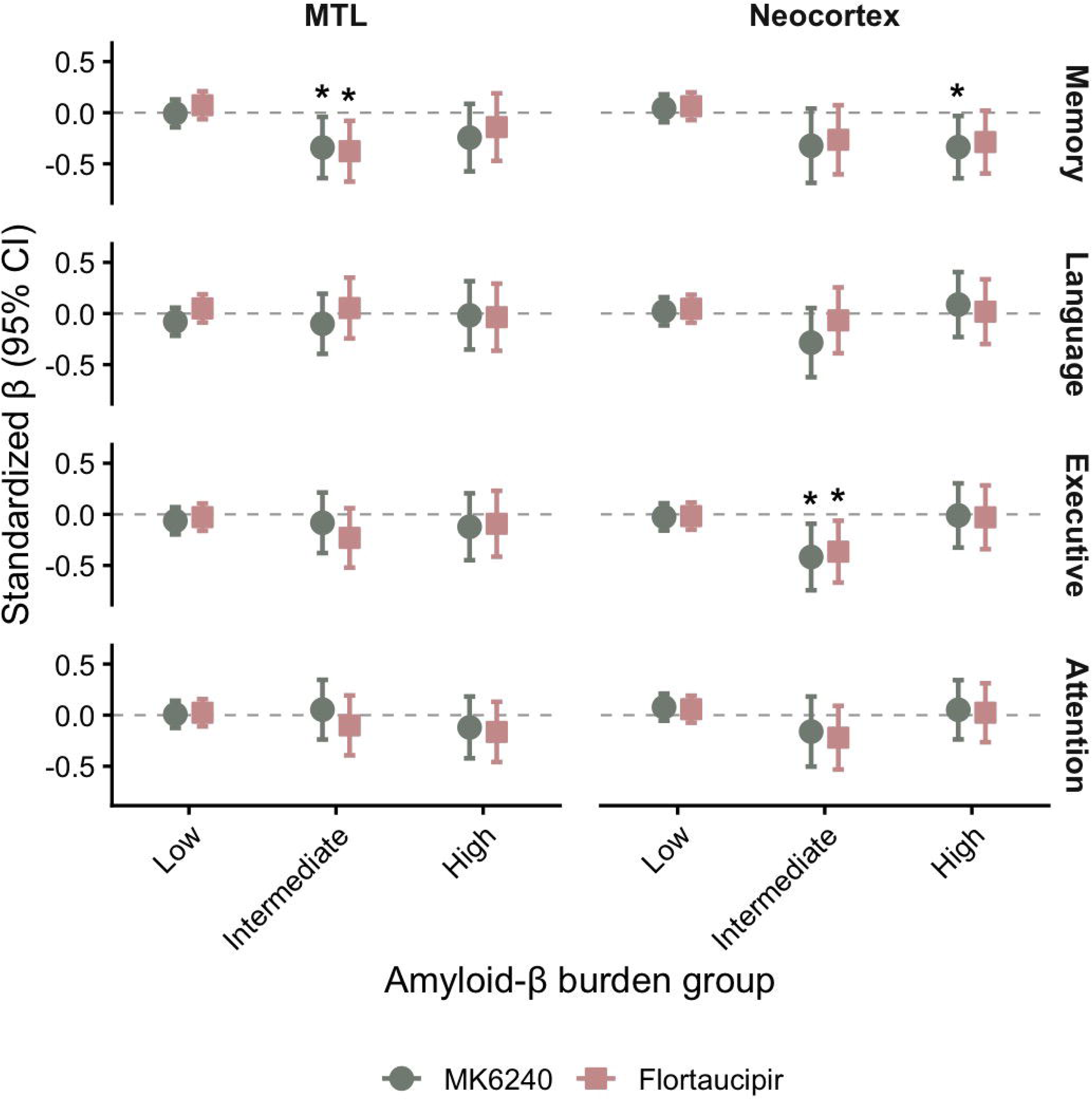
Associations of regional tau-PET SUVR with cognitive performance across amyloid-β burden groups. Standardized regression coefficients (β) and 95% confidence intervals are shown for the associations of medial temporal and neocortical tau-PET SUVR with memory, language, attention, and executive function composite scores, stratified by low, intermediate, and high amyloid-β centiloid burden. Negative coefficients indicate that greater regional tau burden was associated with poorer cognitive performance. Linear models were fitted separately for [¹□F]MK6240 and [¹□F]Flortaucipir and adjusted for age, sex, and education. Nominal p-values are shown. *p<0.05. No associations survived FDR correction. MTL, medial temporal lobe.

After Aβ adjustment, only the [¹□F]flortaucipir–attention clusters remained significant (0.3% of voxels); neither tracer showed significant memory, language, or executive associations.

### Tau-PET burden differs across the SOCI spectrum

In demographic-adjusted analyses, SOCI stage was associated with all regional tau-PET measures except [¹□F]flortaucipir MTL SUVR (ANCOVA p=0.20; **Figure 5A**). Tukey-adjusted comparisons showed greater tau burden in SOCI-3 than all other stages across all [¹□F]MK6240 measures and both [¹□F]flortaucipir SEOT measures (all p<0.05). For [¹□F]flortaucipir neocortical SUVR, SOCI-3 differed only from SOCI-1 (p=0.018). No differences were detected among SOCI-0, SOCI-1, and SOCI-2. Consistent with ROI findings, voxelwise contrasts against SOCI-0 were significant only for SOCI-3 with both tracers, with more extensive differences for [¹□F]MK6240 (**Figure 5B**).

**Figure 5.**
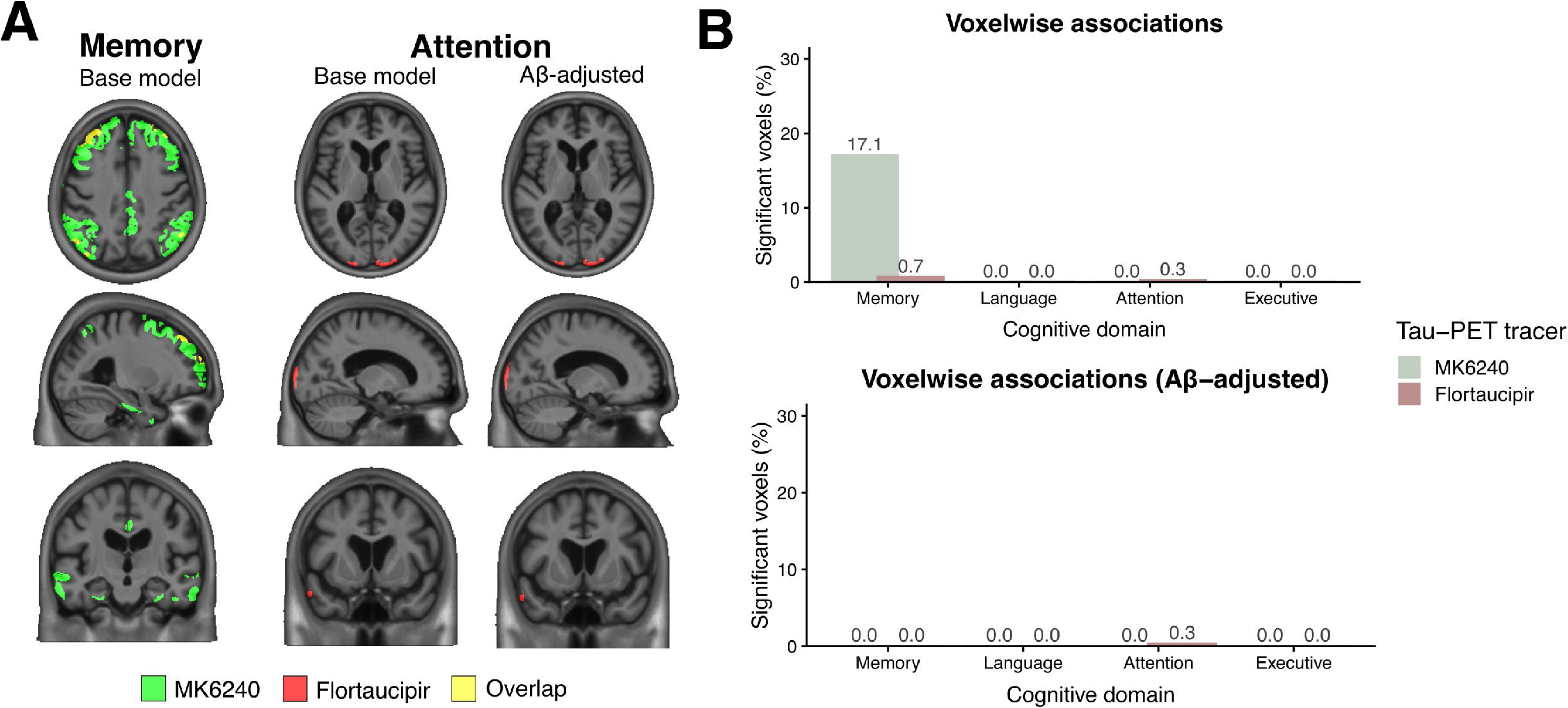
Differential voxelwise associations of [¹□F]MK6240 and [¹□F]Flortaucipir uptake with cognitive performance. **(A)** Cortical voxels showing significant associations between greater tau-PET uptake and poorer performance on the memory and attention composites. Base models were adjusted for age, sex, and education; Aβ-adjusted models were additionally adjusted for centiloids. Only domains with significant associations for at least one tracer are displayed. **(B)** Percentage of cortical voxels showing significant associations for each tracer and cognitive domain in the base and Aβ-adjusted models. Percentages were calculated relative to the total number of voxels within a Braak composite mask. Statistical significance was defined as FDR-corrected p<0.05. Aβ, amyloid-β; FDR, false discovery rate.

**Figure 6.**
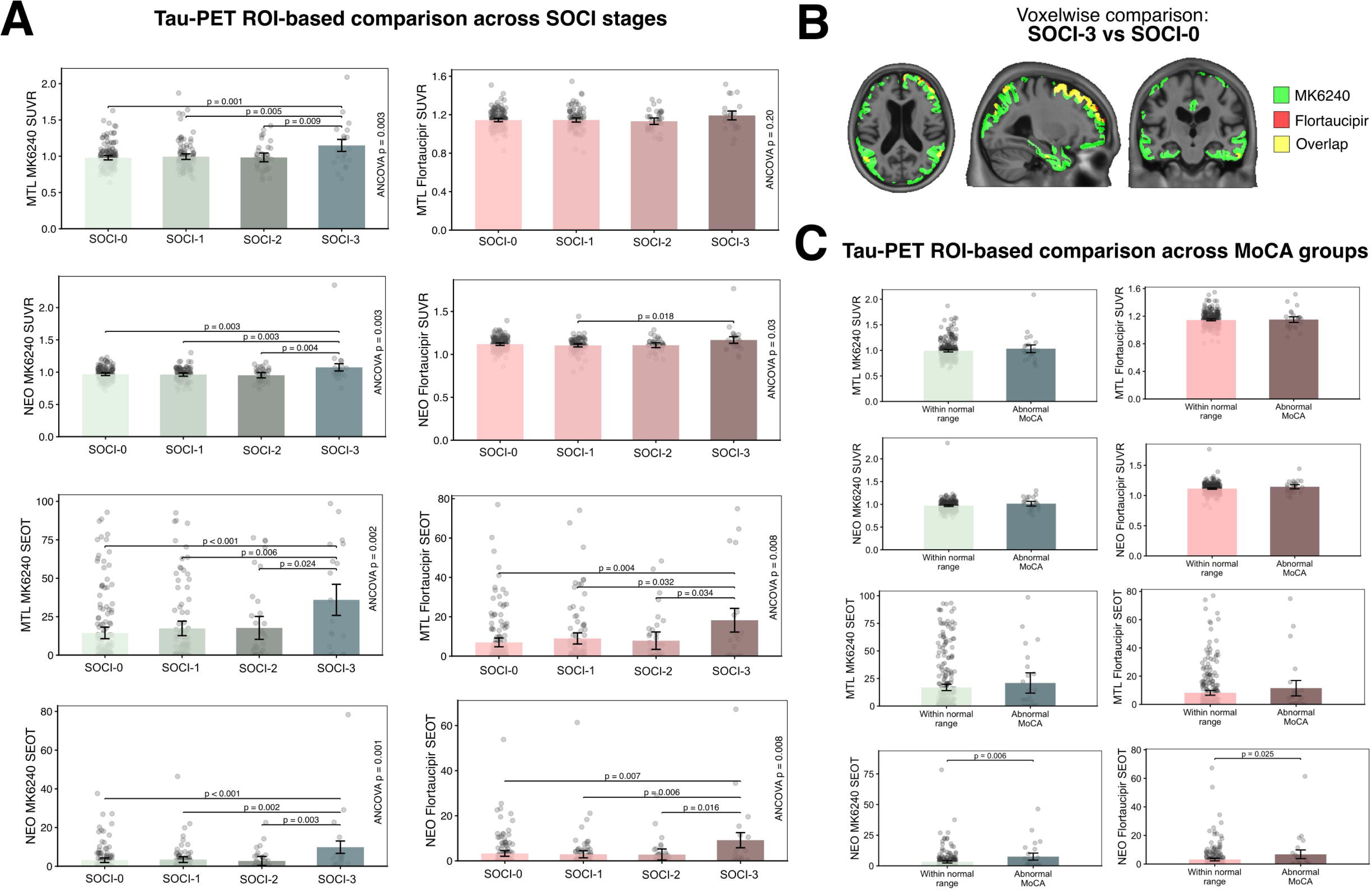
Comparison of regional tau-PET burden across stages of SOCI. **(A)** Regional tau-PET measures across SOCI groups. **(B)** Voxelwise differences between the SOCI-3 and SOCI-0 groups. Maps display voxels surviving FDR correction (*p*<0.05). Other SOCI group contrasts were not significant. **(C)** Regional tau-PET measures according to MoCA classification. No voxelwise differences between MoCA groups were significant. Points represent individual participants; bars and error bars represent estimated marginal means and 95% confidence intervals. Overall group differences were assessed using Type II ANCOVA F-tests. Displayed p-values were obtained from two-sided *t*-tests of estimated marginal means, with Tukey adjustment. All models were adjusted for age, sex, and education. MoCA, Montreal Cognitive Assessment; MTL, medial temporal lobe; NEO, neocortex; SEOT, spatial extent of tau; SUVR, standardised uptake value ratio.

After CL adjustment, overall SOCI-stage differences remained significant only for [¹□F]MK6240 neocortical SEOT (ANCOVA p=0.040) and SUVR (ANCOVA p=0.044; **Supplementary Figure 3A**). Tukey-adjusted comparisons showed greater neocortical SEOT in SOCI-3 than all other stages (p=0.032–0.047), and greater neocortical SUVR than SOCI-1 (p=0.046) and SOCI-2 (p=0.037). Voxelwise differences remained significant only for [¹□F]MK6240 between SOCI-3 and SOCI-0 (**Supplementary Figure 3B**).

Participants with abnormal MoCA (n=25) had greater demographic-adjusted neocortical SEOT than those with normal scores (n=272) for both [¹□F]MK6240 (p=0.006) and [¹□F]flortaucipir (p=0.025; **Figure 5C**). Neither neocortical SUVR nor MTL measures differed significantly. After CL adjustment, only the [¹□F]MK6240 neocortical SEOT difference remained significant (p=0.031; **Supplementary Figure 3C**). No voxelwise differences were significant with either adjustment.

In sensitivity analyses restricted to the entorhinal cortex, SOCI stage was associated with demographic-adjusted [¹□F]MK6240 SUVR (ANCOVA p=0.004) and SEOT (p=0.005), and [¹□F]flortaucipir SEOT (p=0.008), but not SUVR (p=0.11; **Supplementary Figure 4A**). For each significant measure, SOCI-3 had greater tau burden than all other stages in Tukey-adjusted comparisons (all p<0.05). After CL adjustment, overall differences remained significant for SEOT with both tracers (both ANCOVA p=0.04; **Supplementary Figure 4B**). Pairwise differences persisted between SOCI-3 and SOCI-0 for both tracers ([¹□F]MK6240: p=0.03; [¹□F]flortaucipir: p=0.02), and between SOCI-3 and SOCI-2 for [¹□F]MK6240 (p=0.046). No entorhinal measures differed significantly between MoCA groups with either adjustment (**Supplementary Figure 4C-D**).

## Discussion

This cross-sectional, head-to-head study examined associations of [¹□F]flortaucipir and [¹□F]MK6240 measures with subtle objective cognitive deficits in CU individuals. Both tracers showed modest cognitive associations, but [¹□F]MK6240 demonstrated more spatially extensive tau-SOCI associations and more frequent significant differences in uptake between SOCI stages. SEOT complemented SUVR through additional and stronger cognitive associations and more frequent significant differences between SOCI stages. These findings extend evidence that tau-PET captures subtle cognitive variation early in AD, potentially informing participant characterisation and outcome assessment in preventive trials.^12^

Our results align with cross-sectional evidence linking greater tau-PET burden to poorer cognitive performance in CU individuals. Much of this literature has focused on memory, with poorer performance associated with greater MTL tau burden,^8–10^ a pattern also observed in our study. Beyond MTL associations, we found that poorer memory performance was also associated with greater neocortical tau burden, consistent with evidence from across the broader AD continuum.^35^ Emerging evidence further links subtle domain-specific deficits to distinct spatial patterns of tau accumulation in preclinical AD.^11,14^ Our nominal association between poorer executive function and greater neocortical SEOT is consistent with links between non-amnestic deficits and broader neocortical tau accumulation in CU individuals.^11,14^ Conversely, voxelwise [¹□F]flortaucipir uptake was associated with attention in a small superficial occipital cluster. Its proximity to the meninges and skull warrants caution because extracerebral spill-in may explain the association rather than cortical tau.^36,37^ Taken together, our findings reinforce the broader tau-PET literature by showing convergent associations between tau-PET uptake and subtle, domain-specific cognitive performance across two tracers, while also revealing potentially tracer-specific association patterns.

Beyond tracer-related differences, tau–cognition associations also varied across the preclinical AD continuum. No significant associations emerged in CL_LOW_, suggesting that associations at higher Aβ levels may preferentially reflect AD-related tau pathology. Consistent with this interpretation, MTL tau-PET signal in Aβ− CU individuals has been associated with age but not age-adjusted objective cognition.^38^ Tau-cognition associations involved both MTL and neocortical ROIs in CL_INT_ but only neocortical tau in CL_HIGH_. Although none survived FDR correction, this pattern suggests that regional tau correlates of SOCI may differ by Aβ burden. Neocortical associations in CL_INT_ and CL_HIGH_ may reflect Aβ-associated tau spread beyond the MTL, consistent with longitudinal evidence that antecedent Aβ accumulation predicts subsequent tau accumulation, which relates to cognitive decline in CU individuals.^10^ Beyond supporting AD progression models, these findings may inform preventive trials stratifying CU participants by Aβ burden, such as AHEAD 3-45.^12^ Although grouping strategies may differ across trials, our results suggest that regional tau-PET correlates of subtle cognitive variation may vary across Aβ-defined populations. Nevertheless, no subgroup associations survived FDR correction, so these findings remain hypothesis-generating and require validation in larger longitudinal samples.

Although both tracers captured SOCI, only [¹□F]MK6240 showed significant SUVR-based ROI associations with cognitive composites. It also showed broader voxelwise associations with memory, often affected early in typical AD,^39^ and more extensive voxelwise and regional uptake differences across SOCI groups. These findings align with [¹□F]MK6240’s greater affinity and sensitivity for tau neurofibrillary tangles (NFT)^40^ and head-to-head evidence suggesting earlier detection of biologically meaningful tau burden than [¹□F]flortaucipir.^13^ Neuropathological reports also indicate limited [¹□F]flortaucipir sensitivity to low-density MTL tau, particularly without advanced Aβ pathology.^41^ These characteristics may explain broader [¹□F]MK6240 associations with subtle clinical variation here and with other transitional decline features, including mild behavioral impairment.^42^ Despite more frequent significant associations with [¹□F]MK6240, between-tracer ROI effect-size differences did not survive FDR correction. Restricted tau and cognitive ranges in this CU sample may have limited detection of these differences. Broader [¹□F]MK6240 associations therefore do not establish superior clinical performance; implications for individual-level detection, prognosis, and clinical decisions remain uncertain in preclinical AD. Longitudinal studies must determine whether these cross-sectional differences confer meaningful advantages in detecting and predicting the earliest cognitive manifestations of tau pathology.

Beyond tracer comparisons, we examined whether tau-PET metrics influenced SOCI associations within tracers. Both metrics captured cognitive associations, with SEOT performing at least comparably to SUVR and identifying additional significant associations in selected analyses. Notably, higher neocortical SEOT in participants with mildly impaired MoCA scores accords with evidence that MoCA declines as tau extends into the neocortex,^43^ suggesting that SEOT may capture the earliest cognitively detectable phase of this spread. Supporting this interpretation, extent-based measures capture greater interindividual variability at low tau levels and may better reflect early regional tau propagation in AD.^14^ Although SEOT advantages were not consistent across cognitive outcomes, selected associations were stronger and no formal comparison favoured SUVR. Adding SEOT may therefore modestly improve sensitivity to early cognitive differences. This aligns with evidence that tau extent and load differ in their associations with downstream cognitive and neurodegenerative changes, providing potentially complementary information across the AD continuum.^15,44^ Selective advantages may reflect the limited tau burden and cognitive variation expected in CU individuals. Whether this additional sensitivity benefits participant enrichment or target-engagement assessment in preventive trials remains uncertain, although these applications have been proposed.^45^

Beyond continuous cognitive measures, we examined tau-PET differences across SOCI stages adapted from validated actuarial obj-SCD and MCI criteria.^29–31^ Tau-PET burden was comparable across SOCI-0, SOCI-1, and SOCI-2 but higher in SOCI-3 across several measures. These findings provide biomarker support for Jak/Bondi actuarial MCI criteria, complementing evidence of improved diagnostic precision, stronger AD biomarker associations, fewer false positives, and better identification of individuals at risk of progression.^46,47^ Importantly, all participants had global CDR=0 and were classified as CU after extensive evaluation by dementia specialists. Greater tau-PET burden in those meeting actuarial MCI criteria therefore suggests that these criteria may detect biologically meaningful cognitive changes before clinically apparent impairment.

By contrast, SOCI-2 participants (Edmonds/Thomas-like actuarial obj-SCD) did not differ in tau-PET burden from SOCI-0 or SOCI-1. This aligns with reports of no cross-sectional plasma phosphorylated tau 181 or Aβ-PET differences between actuarial obj-SCD and non-impaired controls,^3,4^ but contrasts with higher Braak III–IV [¹□F]flortaucipir SUVR reported in actuarial obj-SCD versus controls without obj-SCD.^5^ That study applied Thomas and colleagues’ extension of Edmonds’ criteria,^5,30^ using different neuropsychological tests and memory-process indices unavailable in UDSv3. Even within the same domain, tests differ in cognitive demands and regional tau-PET associations; substitutions may therefore affect both classification and the tau-related cognitive changes captured by each framework.^48^ Moreover, cross-sectional assessment may limit sensitivity to transitional cognitive change, as SOCI definitions incorporating within-person decline have shown tau-PET associations. Nevertheless, longitudinal measures do not necessarily yield stronger associations with tau pathology.^49^ In a neuropathological study of individuals who remained CU near death, Braak NFT stage alone was not associated with antemortem longitudinal trajectories in episodic memory, executive function, or global cognition.^50^ Thus, longitudinal ascertainment may therefore improve SOCI construct validity without ensuring direct correspondence with tau burden. These findings highlight the need to compare heterogeneous SOCI definitions^2^ for biomarker and prognostic validity. Extending criteria to broader batteries and validating alternative neuropsychological measures may clarify which psychometric definitions best characterize SOCI within Alzheimer’s Association clinical stage 2.^1^

Study strengths include a unique, substantial sample of well-characterized CU individuals imaged with two widely used tau-PET tracers. However, without gold-standard neuropathological evaluation, we cannot directly establish which tracer more accurately reflects biologically relevant NFT pathology. The multicenter design reduces single-site dependence, but exclusively North American recruitment warrants replication in more diverse populations and settings to establish generalizability. Another limitation is the cross-sectional operationalization of the SOCI. Because longitudinal data from the HEAD cohort are not yet extensively available, we could not incorporate intra-individual longitudinal cognitive change, as adopted in other work^49^ and in the Alzheimer’s Association workgroup framework.^1^ Moreover, although SOCI criteria were adapted from validated approaches,^29–31^ battery differences prevented exact implementation and may have influenced classification and biomarker associations. Despite these limitations, our findings extend biomarker characterization of a widely used SOCI operationalization.^2–6^

In sum, both [¹□F]MK6240 and [¹□F]Flortaucipir captured subtle, domain-specific cognitive variation in CU individuals, with spatially broader associations for [¹□F]MK6240. SEOT complemented SUVR in detecting early tau-cognition relationships. Higher tau burden in SOCI-3 but not SOCI-2 suggests that actuarial MCI criteria may identify biologically meaningful cognitive deficits before clinical diagnosis.

## Author Contributions

ACM and PRN designed or conceptualized the study. ACM, LT, FZL, TC, CT, NR, GP, PCLF, BB, JFA, LA, ER, MSM, RM, AR, BP, JM, BAG, VJL, HO, DNSM, JF, SB, TAP, and PRN had major roles in the acquisition of data. ACM, EA, and PRN analysed or interpreted the data. ACM drafted the manuscript. All revised the manuscript for intellectual content.

## Acknowledgements

This work would not have been possible without the generosity of all the individuals who volunteered to participate in the HEAD study. We are grateful to all study participants, their families, and caregivers for their time, trust, and commitment to this research. We extend our appreciation to the clinical and research staff at the participating sites (University of Pittsburgh, Lawrence Berkeley National Laboratory, Houston Methodist Research Institute, Washington University in St Louis, Mayo Clinic, Brown University, University of California San Francisco, and McGill University) for their dedication to participant recruitment, study coordination, and data collection. We also thank all referring clinicians and community partners for their support of recruitment at all sites. We gratefully acknowledge the National Alzheimer’s Coordinating Center for permission to use the Uniform Data Set in this study. We acknowledge the Laboratory of Neuroimaging for providing the imaging and data repository, and the National Centralized Repository for Alzheimer’s Disease and Related Dementias for providing the biological sample repository and APOE genotyping in this study. Finally, we extend our thanks to Lantheus/Cerveau Technologies and Lilly/Avid Radiopharmaceuticals for enabling use of the [^18^F]MK6240 and [^18^F]Flortaucipir radiotracers, respectively, in this comparison study. The HEAD study is funded by the National Institute on Aging (R01AG073267). This manuscript is the result of funding in whole or in part by the National Institutes of Health (NIH). It is subject to the NIH Public Access Policy. Through acceptance of this federal funding, NIH has been given a right to make this manuscript publicly available in PubMed Central upon the Official Date of Publication, as defined by NIH. ACM is supported by the Alzheimer Society of Canada Doctoral Award. IB is supported by the Coordenação de Aperfeiçoamento de Pessoal de Nível Superior (88881.218211/2025-01). GP receives financial support from the Alzheimer’s Association (24AARFD-1243899). BB is supported by the Alzheimer’s Association (AARFD-22–974627). PCLF is supported by the Alzheimer’s Association (AARFD-22–923814). PR-N receives funding from the Weston Brain Institute, Canadian Institutes of Health Research Fonds de Recherche du Québec – Santé (FRQS), Colin J Adair Charitable Foundation. JF reports grants from the Fondo de Investigaciones Sanitario, Carlos III Health Institute (INT21/00073 and PI23/01786) and the Centro de Investigación Biomédica en Red sobre Enfermedades Neurodegenerativas Program 1, jointly funded by Fondo Europeo de Desarrollo Regional, Unión Europea, Una manera de hacer Europa. JF was also supported by NIH grants (1R01AG056850–01A1; R21AG056974, R01AG061566, 1R01AG081394–01, and 1R61AG066543–01), the Department de Salut de la Generalitat de Catalunya (SLT006/17/00119), and Fundación Tatiana Pérez de Guzmán el Bueno (IIBSP-DOW-2020–151). He was also supported by Horizon 2020–Research and Innovation Framework Programme from the European Union (H2020-SC1-BHC-2018–2020). DNS-M receives research support from the NIH (K23-AG076960). Development of the National Alzheimer’s Coordinating Center Uniform Data Set was funded by the NIH through the National Institute on Aging (Cooperative Agreement U01AG016976). The Laboratory of Neuroimaging is supported by the NIH through the National Institute of Biomedical Imaging and Bioengineering (P41EB015922); the National Centralized Repository for Alzheimer’s Disease and Related Dementias receives government support under a cooperative agreement grant (U24AG021886) awarded by the National Institute on Aging.

## Contributor information

for the HEAD Study: Cristiano S Aguzzoli, Livia Amaral, Guilherme Bauer-Negrini, Bruna Bellaver, Ann D Cohen, Alvaro de Oliveira Franco, Douglas T Leffa, Pamela C L Ferreira, Firoza Z Lussier, Marina S Medeiros, Rayan Mroué, Markley S Oliveira, Tharick A Pascoal, Guilherme Povala, Andréia Rocha, Matheus S Rodrigues, Emma Ruppert, Pampa Saha, Juli Singer, Carolina Soares, Dana L Tudorascu, Raphael L Tuma (Department of Psychiatry, University of Pittsburgh, PA, USA). Suzanne L Baker, Cécile Tissot, Hsin-Yeh Tsai (Lawrence Berkeley National Laboratory, Berkeley, CA, USA). Pedro Rosa-Neto (Peter O’Donnell Jr Brain Institute, University of Texas Southwestern Medical Centre, Dallas, TX, USA). Tevy Chan, Vladimir Fonov, Yasser Iturria-Medina, Wan Lu Jia, Jesse Klostranec, Maxime Montembault, Nesrine Rahmouni, Jenna Stevenson, Karine Provost, Paolo Vitali (McGill University, Montreal, QC, Canada). Selma Avdagic, Emerine Cummings, Brian A Gordon, Danielle Gray, Victoria Tate (Department of Radiology, Washington University in St Louis, St Louis, MO, USA). Joseph C Masdeu and Belen Pascual (Department of Neurology, Houston Methodist Research Institute and Weill Cornell Medicine, Houston, TX, USA). Hwamee Oh and Xiaoyang Hu (Department of Psychiatry and Human Behavior, Brown University, Providence, RI, USA). Val Lowe (Department of Radiology, Mayo Clinic, Rochester, MN, USA). David N Soleimani-Meigooni (Edward and Pearl Fein Memory and Aging Center, University of California San Francisco, San Francisco, CA, USA). Juan Fortea (Sant Pau Memory Unit, IR SANT PAU, Hospital de la Santa Creu i Sant Pau, Barcelona, Spain). Arthur Toga (Mark and Mary Stevens Neuroimaging and Informatics Institute, University of Southern California, CA, USA). Tatiana Foroud (Department of Medical and Molecular Genetics, Indiana University School of Medicine, Indianapolis, IN, USA).

