## Supplementary materials for "Head-to-Head Comparison of Flortaucipir and MK6240 tau-PET to Detect Subtle Objective Cognitive Impairment in Clinically Unimpaired Individuals"

### **Supplementary Table 1. Associations between regional tau-PET SUVR and cognitive performance**

| **Cognitive outcome** | **Tracer** | **Region** | **Standardized β (95% CI)** | **Nominal p** | **FDR p** | **Adjusted R²** | **F statistics** |
| --- | --- | --- | --- | --- | --- | --- | --- |
| Memory | MK6240 | MTL | -0.15 (-0.27 to -0.03) | 0.012 | 0.042 | 0.045 | 4.52 |
|  | MK6240 | Neocortex | -0.13 (-0.25 to -0.02) | 0.021 | 0.042 | 0.042 | 4.25 |
|  | Flortaucipir | MTL | -0.06 (-0.18 to 0.06) | 0.319 | 0.319 | 0.028 | 3.11 |
|  | Flortaucipir | Neocortex | -0.08 (-0.19 to 0.04) | 0.184 | 0.245 | 0.030 | 3.32 |
| Language | MK6240 | MTL | -0.07 (-0.19 to 0.05) | 0.235 | 0.894 | 0.069 | 6.52 |
|  | MK6240 | Neocortex | -0.01 (-0.12 to 0.10) | 0.894 | 0.894 | 0.065 | 6.14 |
|  | Flortaucipir | MTL | 0.03 (-0.08 to 0.15) | 0.583 | 0.894 | 0.066 | 6.22 |
|  | Flortaucipir | Neocortex | 0.02 (-0.10 to 0.13) | 0.774 | 0.894 | 0.065 | 6.16 |
| Attention | MK6240 | MTL | 0.00 (-0.11 to 0.11) | 0.990 | 0.990 | 0.122 | 11.31 |
|  | MK6240 | Neocortex | 0.04 (-0.07 to 0.15) | 0.484 | 0.990 | 0.124 | 11.45 |
|  | Flortaucipir | MTL | -0.03 (-0.14 to 0.09) | 0.645 | 0.990 | 0.123 | 11.37 |
|  | Flortaucipir | Neocortex | 0.01 (-0.10 to 0.11) | 0.931 | 0.990 | 0.122 | 11.31 |
| Executive | MK6240 | MTL | -0.09 (-0.20 to 0.03) | 0.131 | 0.138 | 0.116 | 10.72 |
|  | MK6240 | Neocortex | -0.08 (-0.19 to 0.03) | 0.138 | 0.138 | 0.116 | 10.70 |
|  | Flortaucipir | MTL | -0.09 (-0.20 to 0.03) | 0.134 | 0.138 | 0.116 | 10.71 |
|  | Flortaucipir | Neocortex | -0.09 (-0.20 to 0.01) | 0.089 | 0.138 | 0.118 | 10.90 |

Each row represents a separate linear regression adjusted for age, sex, and years of education. β denotes the standardized regression coefficient for the tau-PET metric. False discovery rate p-values were calculated using the Benjamini-Hochberg procedure across the four tracer-by-region associations within each cognitive domain. The F statistic and adjusted R² describe the complete fitted model. MTL, medial temporal lobe; SEOT, spatial extent of tauopathy; SUVR, standardized uptake value ratio.

### **Supplementary Table 2. Associations between tau-PET SUVR and cognitive performance after additional adjustment for Centiloid**

| **Cognitive outcome** | **Tracer** | **Region** | **Standardized β (95% CI)** | **Nominal p** | **FDR q** | **Adjusted R²** | **F statistics** |
| --- | --- | --- | --- | --- | --- | --- | --- |
| Memory | MK6240 | MTL | -0.07 (-0.20 to 0.06) | 0.286 | 0.571 | 0.065 | 5.11 |
|  | MK6240 | Neocortex | -0.08 (-0.19 to 0.04) | 0.206 | 0.571 | 0.066 | 5.21 |
|  | Flortaucipir | MTL | -0.01 (-0.13 to 0.11) | 0.926 | 0.926 | 0.061 | 4.87 |
|  | Flortaucipir | Neocortex | -0.03 (-0.15 to 0.08) | 0.599 | 0.799 | 0.062 | 4.92 |
| Language | MK6240 | MTL | -0.07 (-0.20 to 0.06) | 0.306 | 0.876 | 0.066 | 5.20 |
|  | MK6240 | Neocortex | 0.00 (-0.11 to 0.12) | 0.947 | 0.947 | 0.063 | 4.97 |
|  | Flortaucipir | MTL | 0.04 (-0.08 to 0.16) | 0.464 | 0.876 | 0.065 | 5.09 |
|  | Flortaucipir | Neocortex | 0.03 (-0.09 to 0.14) | 0.657 | 0.876 | 0.064 | 5.01 |
| Attention | MK6240 | MTL | -0.03 (-0.16 to 0.09) | 0.614 | 0.872 | 0.123 | 9.32 |
|  | MK6240 | Neocortex | 0.02 (-0.09 to 0.14) | 0.697 | 0.872 | 0.123 | 9.29 |
|  | Flortaucipir | MTL | -0.04 (-0.16 to 0.07) | 0.451 | 0.872 | 0.124 | 9.39 |
|  | Flortaucipir | Neocortex | -0.01 (-0.12 to 0.10) | 0.872 | 0.872 | 0.122 | 9.26 |
| Executive | MK6240 | MTL | -0.08 (-0.21 to 0.04) | 0.196 | 0.196 | 0.113 | 8.55 |
|  | MK6240 | Neocortex | -0.08 (-0.19 to 0.04) | 0.194 | 0.196 | 0.113 | 8.55 |
|  | Flortaucipir | MTL | -0.08 (-0.20 to 0.04) | 0.180 | 0.196 | 0.114 | 8.58 |
|  | Flortaucipir | Neocortex | -0.09 (-0.20 to 0.02) | 0.119 | 0.196 | 0.115 | 8.72 |

Each row represents a separate linear regression adjusted for age, sex, years of education, and Centiloid. β denotes the standardized regression coefficient for the tau-PET metric. False discovery rate p-values were calculated using the Benjamini-Hochberg procedure across the four tracer-by-region associations within each cognitive domain. The F statistic and adjusted R² describe the complete fitted model. MTL, medial temporal lobe; SEOT, spatial extent of tauopathy; SUVR, standardized uptake value ratio.

**Supplementary Table 3. Associations between entorhinal tau-PET SUVR and cognitive performance**

| **Cognitive outcome** | **Tracer** | **Standardized β (95% CI)** | **Nominal p** | **FDR p** | **Adjusted R²** | **F statistic** |
| --- | --- | --- | --- | --- | --- | --- |
| Memory | MK6240 | -0.11 (-0.20 to -0.02) | **0.02** | **0.03** | 0.04 | 4.41 |
|  | Flortaucipir | -0.05 (-0.13 to 0.04) | 0.30 | 0.30 | 0.03 | 3.14 |
| Language | MK6240 | -0.05 (-0.12 to 0.03) | 0.23 | 0.46 | 0.07 | 6.53 |
|  | Flortaucipir | 0.01 (-0.06 to 0.08) | 0.75 | 0.75 | 0.07 | 6.17 |
| Attention | MK6240 | -0.01 (-0.09 to 0.07) | 0.80 | 0.80 | 0.12 | 11.33 |
|  | Flortaucipir | -0.03 (-0.11 to 0.05) | 0.41 | 0.80 | 0.12 | 11.51 |
| Executive | MK6240 | -0.07 (-0.17 to 0.02) | 0.13 | 0.25 | 0.12 | 10.74 |
|  | Flortaucipir | -0.05 (-0.14 to 0.04) | 0.26 | 0.26 | 0.11 | 10.44 |

Each row represents a separate linear regression adjusted for age, sex, and years of education. β denotes the regression coefficient for entorhinal SUVR. Benjamini–Hochberg FDR correction was applied across the two tracers within each cognitive domain, separately for each adjustment model. Bold p-values indicate significance at p < 0.05 in the corresponding column, determined before rounding. Adjusted R² and the F statistic describe the complete fitted model. CI, confidence interval; FDR, false discovery rate; SUVR, standardized uptake value ratio .

**Supplementary Table 4. Associations between entorhinal tau-PET SUVR and cognitive performance after additional adjustment for Centiloid**

| **Cognitive outcome** | **Tracer** | **Standardized β (95% CI)** | **Nominal p** | **FDR p** | **Adjusted R²** | **F statistic** |
| --- | --- | --- | --- | --- | --- | --- |
| Memory | MK6240 | -0.06 (-0.16 to 0.04) | 0.22 | 0.45 | 0.07 | 5.19 |
|  | Flortaucipir | -0.01 (-0.10 to 0.08) | 0.86 | 0.86 | 0.06 | 4.87 |
| Language | MK6240 | -0.04 (-0.12 to 0.04) | 0.30 | 0.59 | 0.07 | 5.21 |
|  | Flortaucipir | 0.02 (-0.06 to 0.09) | 0.63 | 0.63 | 0.06 | 5.02 |
| Attention | MK6240 | -0.03 (-0.12 to 0.06) | 0.49 | 0.49 | 0.12 | 9.37 |
|  | Flortaucipir | -0.05 (-0.13 to 0.03) | 0.27 | 0.49 | 0.13 | 9.55 |
| Executive | MK6240 | -0.07 (-0.17 to 0.03) | 0.18 | 0.33 | 0.11 | 8.57 |
|  | Flortaucipir | -0.05 (-0.14 to 0.05) | 0.33 | 0.33 | 0.11 | 8.38 |

Each row represents a separate linear regression adjusted for age, sex, and years of education, with additional adjustment for centiloid. β denotes the regression coefficient for entorhinal SUVR. Benjamini–Hochberg FDR correction was applied across the two tracers within each cognitive domain. Bold p-values indicate significance at p < 0.05 in the corresponding column, determined before rounding. Adjusted R² and the F statistic describe the complete fitted model. CI, confidence interval; FDR, false discovery rate; SUVR, standardized uptake value ratio .

**Supplementary Table 5. Associations between tau-PET spatial extent and cognitive performance**

| **Cognitive outcome** | **Tracer** | **Region** | **Standardized β (95% CI)** | **Nominal p** | **FDR p** | **Adjusted R²** | **F statistics** |
| --- | --- | --- | --- | --- | --- | --- | --- |
| Memory | MK6240 | MTL | -0.16 (-0.28 to -0.04) | 0.010 | 0.010 | 0.046 | 4.59 |
|  | MK6240 | Neocortex | -0.18 (-0.29 to -0.07) | 0.002 | 0.003 | 0.058 | 5.52 |
|  | Flortaucipir | MTL | -0.16 (-0.28 to -0.04) | 0.008 | 0.010 | 0.047 | 4.69 |
|  | Flortaucipir | Neocortex | -0.19 (-0.31 to -0.08) | 7.29 x 10^-4^ | 0.003 | 0.062 | 5.88 |
| Language | MK6240 | MTL | -0.09 (-0.20 to 0.03) | 0.148 | 0.592 | 0.072 | 6.71 |
|  | MK6240 | Neocortex | -0.03 (-0.14 to 0.08) | 0.560 | 0.766 | 0.066 | 6.23 |
|  | Flortaucipir | MTL | -0.02 (-0.14 to 0.10) | 0.766 | 0.766 | 0.065 | 6.16 |
|  | Flortaucipir | Neocortex | -0.02 (-0.14 to 0.09) | 0.674 | 0.766 | 0.065 | 6.19 |
| Attention | MK6240 | MTL | -0.05 (-0.16 to 0.07) | 0.429 | 0.858 | 0.124 | 11.49 |
|  | MK6240 | Neocortex | -0.01 (-0.12 to 0.09) | 0.802 | 0.869 | 0.122 | 11.33 |
|  | Flortaucipir | MTL | -0.06 (-0.18 to 0.05) | 0.283 | 0.858 | 0.126 | 11.65 |
|  | Flortaucipir | Neocortex | -0.01 (-0.12 to 0.10) | 0.869 | 0.869 | 0.122 | 11.32 |
| Executive | MK6240 | MTL | -0.13 (-0.24 to -0.01) | 0.027 | 0.055 | 0.128 | 11.76 |
|  | MK6240 | Neocortex | -0.11 (-0.22 to 0.00) | 0.048 | 0.064 | 0.129 | 11.88 |
|  | Flortaucipir | MTL | -0.13 (-0.25 to -0.02) | 0.022 | 0.055 | 0.133 | 12.23 |
|  | Flortaucipir | Neocortex | -0.07 (-0.18 to 0.03) | 0.176 | 0.176 | 0.123 | 11.29 |

Each row represents a separate linear regression adjusted for age, sex, and years of education. β denotes the standardized regression coefficient for the tau-PET metric. False discovery rate p-values were calculated using the Benjamini-Hochberg procedure across the four tracer-by-region associations within each cognitive domain. The F statistic and adjusted R² describe the complete fitted model. MTL, medial temporal lobe; SEOT, spatial extent of tauopathy; SUVR, standardized uptake value ratio.

### **Supplementary Table 6. Associations between tau-PET spatial extent and cognitive performance after additional adjustment for Centiloid**

| **Cognitive outcome** | **Tracer** | **Region** | **Standardized β (95% CI)** | **Nominal p** | **FDR p** | **Adjusted R²** | **F statistics** |
| --- | --- | --- | --- | --- | --- | --- | --- |
| Memory | MK6240 | MTL | -0.08 (-0.21 to 0.05) | 0.248 | 0.248 | 0.066 | 5.15 |
|  | MK6240 | Neocortex | -0.13 (-0.25 to -0.01) | 0.035 | 0.070 | 0.076 | 5.84 |
|  | Flortaucipir | MTL | -0.12 (-0.24 to 0.00) | 0.059 | 0.079 | 0.073 | 5.64 |
|  | Flortaucipir | Neocortex | -0.15 (-0.27 to -0.04) | 0.008 | 0.033 | 0.084 | 6.40 |
| Language | MK6240 | MTL | -0.09 (-0.22 to 0.04) | 0.189 | 0.756 | 0.068 | 5.35 |
|  | MK6240 | Neocortex | -0.02 (-0.14 to 0.10) | 0.692 | 0.876 | 0.063 | 5.01 |
|  | Flortaucipir | MTL | -0.01 (-0.13 to 0.11) | 0.876 | 0.876 | 0.063 | 4.98 |
|  | Flortaucipir | Neocortex | -0.02 (-0.13 to 0.10) | 0.778 | 0.876 | 0.063 | 4.99 |
| Attention | MK6240 | MTL | -0.09 (-0.22 to 0.04) | 0.164 | 0.348 | 0.128 | 9.71 |
|  | MK6240 | Neocortex | -0.04 (-0.15 to 0.08) | 0.516 | 0.670 | 0.124 | 9.36 |
|  | Flortaucipir | MTL | -0.08 (-0.20 to 0.04) | 0.174 | 0.348 | 0.128 | 9.69 |
|  | Flortaucipir | Neocortex | -0.02 (-0.14 to 0.09) | 0.670 | 0.670 | 0.123 | 9.30 |
| Executive | MK6240 | MTL | -0.13 (-0.26 to -0.01) | 0.038 | 0.076 | 0.121 | 9.15 |
|  | MK6240 | Neocortex | -0.11 (-0.22 to 0.01) | 0.069 | 0.092 | 0.118 | 8.93 |
|  | Flortaucipir | MTL | -0.13 (-0.25 to -0.01) | 0.030 | 0.076 | 0.122 | 9.25 |
|  | Flortaucipir | Neocortex | -0.07 (-0.18 to 0.04) | 0.231 | 0.231 | 0.112 | 8.49 |

Each row represents a separate linear regression adjusted for age, sex, years of education, and Centiloid. β denotes the standardized regression coefficient for the tau-PET metric. False discovery rate q values were calculated using the Benjamini-Hochberg procedure across the four tracer-by-region associations within each cognitive domain. The F statistic and adjusted R² describe the complete fitted model. MTL, medial temporal lobe; SEOT, spatial extent of tauopathy; SUVR, standardized uptake value ratio.

**Supplementary Figure 1. Masks for the regions-of-interest and voxelwise analysis template.**

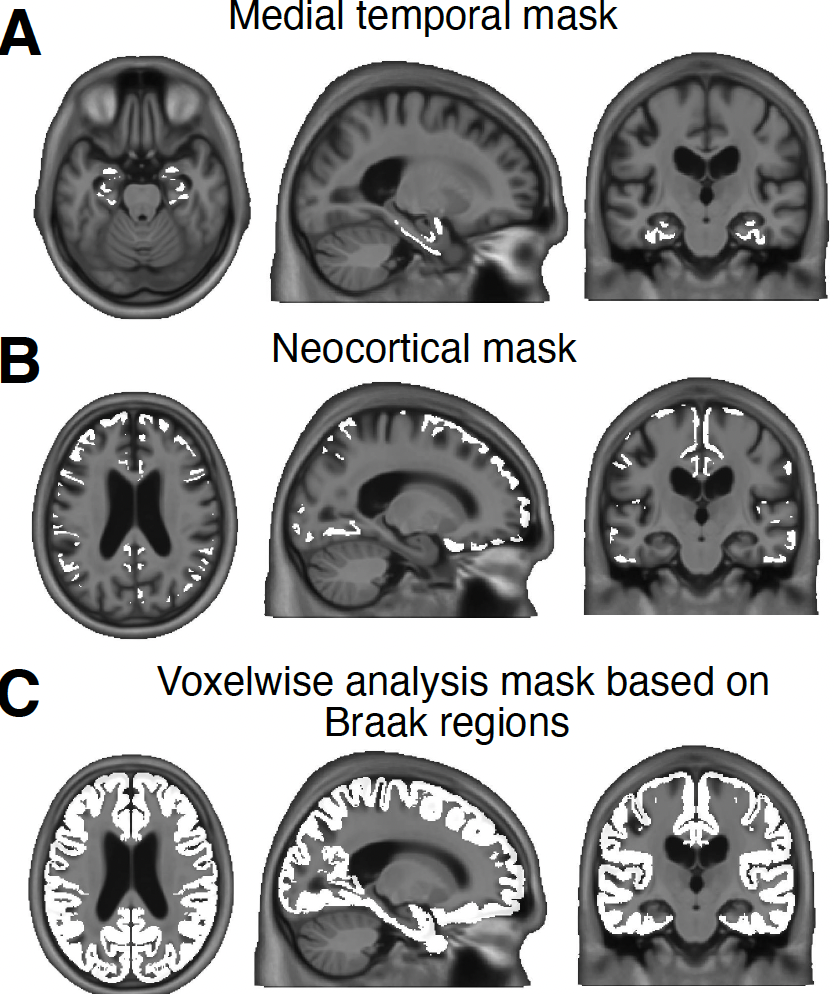

**Supplementary Figure 2.** **Comparison of SEOT and SUVR associations with cognition within each tau-PET tracer.** Standardized regression coefficients were compared between SEOT and SUVR for [^18^F]MK6240 (top) and [^18^F]Flortaucipir (bottom) in the medial temporal lobe (MTL; left) and neocortex (right). Models were adjusted for age, sex, and years of education. Bars and black points indicate the observed coefficient difference (βSEOT − βSUVR), and horizontal lines represent 95% percentile confidence intervals derived from 5,000 paired participant-level bootstrap resamples. Negative differences indicate more negative associations for SEOT (darker shades), whereas positive differences indicate more negative associations for SUVR (lighter shades). Nominal and Benjamini–Hochberg FDR-adjusted p values are shown; correction was applied separately for each cognitive outcome across four comparisons (two tracers × two ROIs). Bold values indicate p < 0.05 in the corresponding column. FDR, false discovery rate; SEOT, spatial extent of tau; SUVR, standardized uptake value ratio.
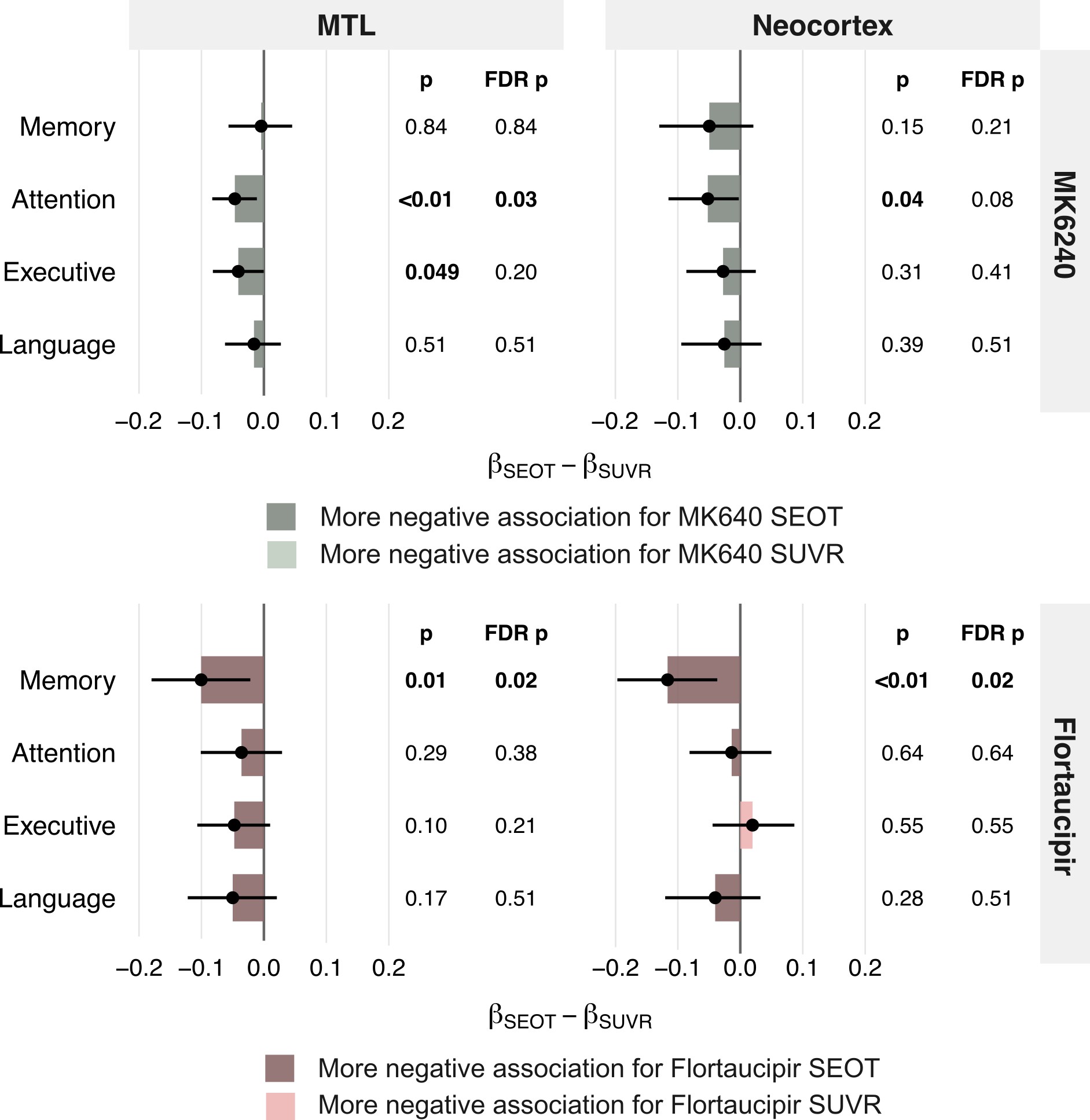

**Supplementary Figure 3. Comparison of regional tau-PET burden across stages of SOCI after adjustment for amyloid-β burden.** **(A)** Regional tau-PET measures across SOCI groups. **(B)** Voxelwise differences between the SOCI-3 and SOCI-0 groups, in which only [^18^F]MK6240 showed significant clusters. Maps display voxels surviving FDR correction (*p*<0.05). Other SOCI group contrasts were not significant. **(C)** Regional tau-PET measures according to MoCA classification. No voxelwise differences between MoCA groups were significant. Points represent individual participants; bars and error bars represent estimated marginal means and 95% confidence intervals. Overall group differences were assessed using Type II ANCOVA F-tests. Displayed p-values were obtained from two-sided *t*-tests of estimated marginal means, with Tukey adjustment. All models were adjusted for age, sex, education, and amyloid-β burden. MoCA, Montreal Cognitive Assessment; MTL, medial temporal lobe; NEO, neocortex; SEOT, spatial extent of tau; SOCI, subtle objective cognitive impairment; SUVR, standardized uptake value ratio.

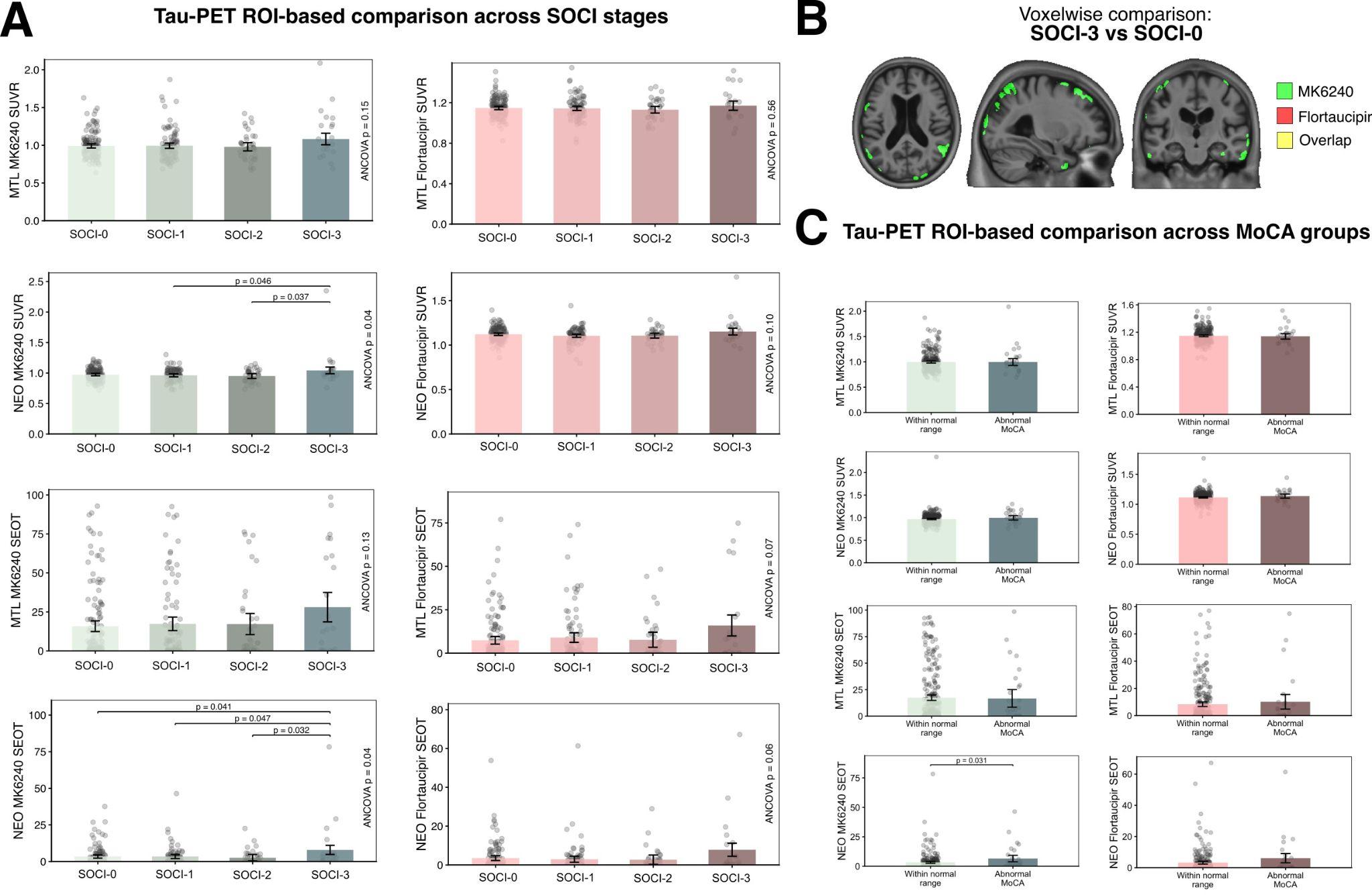

**Supplementary Figure 4. Comparison of entorhinal tau-PET burden across SOCI stages and MoCA groups.** Entorhinal SUVR and SEOT for MK6240 and flortaucipir are shown across SOCI stages **(A)** after adjustment for age, sex, and education and **(B)** after additional adjustment for amyloid-β burden measured in Centiloids. **(C)** and **(D)** respectively show tau-PET measure comparisons between MoCA groups without and with additional amyloid-β adjustment. Points represent individual participants; bars and error bars represent estimated marginal means and 95% confidence intervals. Overall group differences were assessed using Type II ANCOVA F-tests. Brackets indicate significant pairwise comparisons based on two-sided *t*-tests of estimated marginal means, with Tukey adjustment for SOCI comparisons. No MoCA group comparisons were significant. SEOT, spatial extent of tauopathy; SOCI, subtle objective cognitive impairment; SUVR, standardised uptake value ratio; MoCA, Montreal Cognitive Assessment.
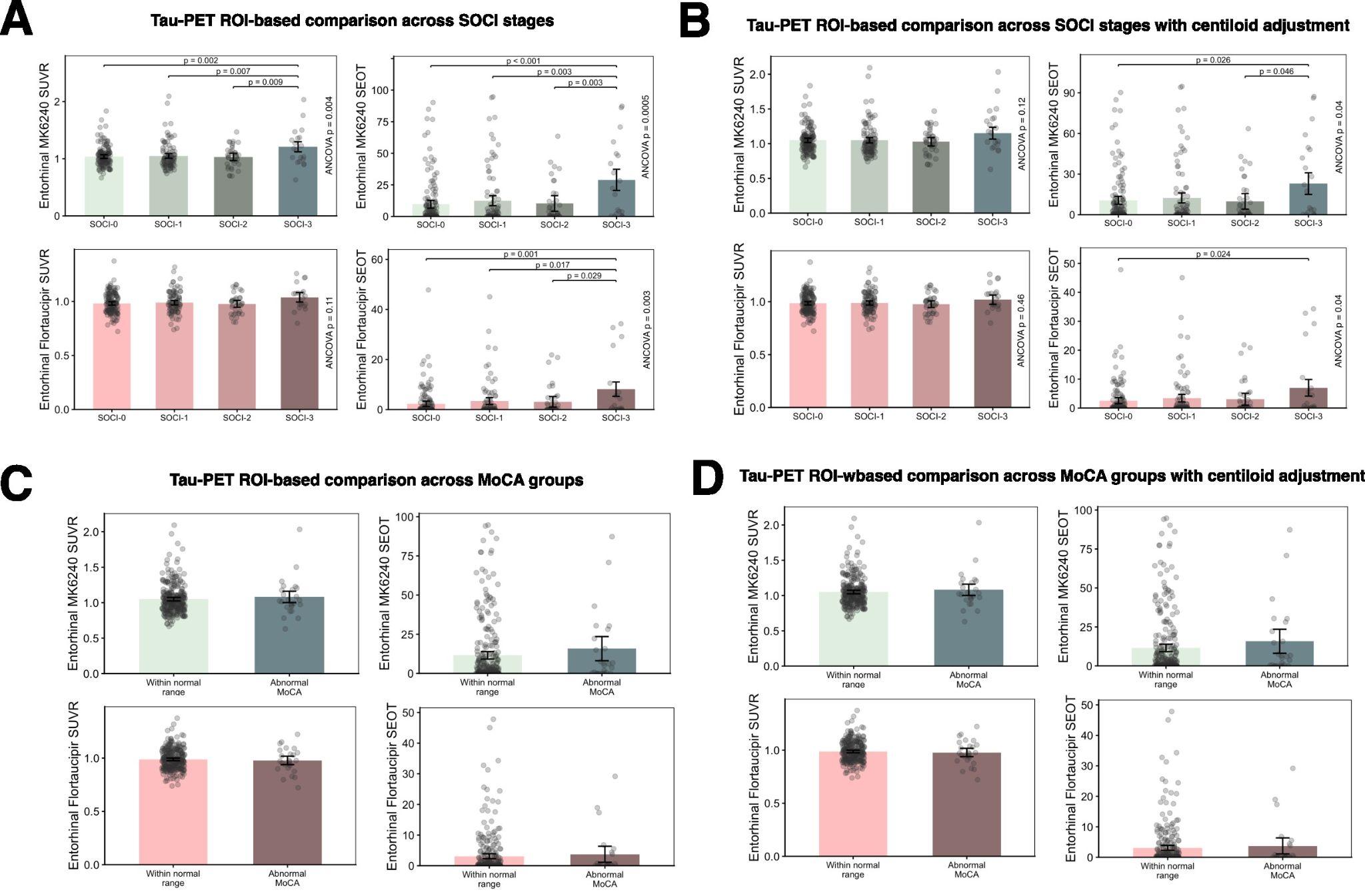
